# Genomic epidemiology reconstructs the pre-detection transmission and establishment of chikungunya virus in Guangdong, China

**DOI:** 10.64898/2026.11.04.26367289

**Authors:** Xiaofang Peng, Zhiyuan Chen, Penghui Jia, Chang Zhang, Meng Zhang, Yuwen Zhong, Andi Sun, Shen Huang, Wenzhi Li, Xin Zhang, Jun Cai, Zhe Liu, Junteng Luo, Juan Su, Changyun Sun, Qingzi Yu, Qiqi Tan, Zhonghua Zheng, Lirong Zou, Jianfeng He, Nuno R. Faria, Baisheng Li, Hongjie Yu

## Abstract

In 2025, a chikungunya outbreak in southern China resulted in more than 25,000 confirmed cases, but its origins and transmission history remain unresolved. As part of the public health investigation, we generated 1,264 near-complete chikungunya virus (CHIKV) genomes from Guangdong Province and integrated genomic, epidemiological, entomological, and human mobility data to reconstruct the emergence and spread of this outbreak. Our analyses traced the outbreak to a single dominant introduction of a Réunion-associated East/Central/South African (ECSA) virus lineage and dated the origin of the sampled Guangdong outbreak lineage from late April to early May 2025. The interval between the estimated origin of the outbreak lineage and the first reported case was consistent with a period of undetected local transmission and exposed limitations in passive surveillance for early arboviral outbreak detection. Spatiotemporal reconstruction showed two distinct epidemic waves with contrasting transmission dynamics. The Foshan wave involved frequent medium- to long-distance viral movement, while the Jiangmen wave remained relatively localized. Together, these findings reconstruct the early establishment and spatially heterogeneous spread of the largest documented CHIKV epidemic in southern China to date and demonstrate the necessity of expanding active genomic surveillance in areas at the fringe of CHIKV endemic transmission for timely detecting and characterizing emerging epidemics.

## Introduction

Chikungunya virus (CHIKV) is a mosquito-borne, single-stranded, positive-sense RNA virus of approximately 11.8 kb that is transmitted in human-amplified cycles by *Aedes aegypti* and *Ae. albopictus*. Human infection typically causes an acute febrile illness, often accompanied by debilitating arthralgia that may persist for months^1^. CHIKV belongs to the *Togaviridae* family and can be classified into three major phylogenetic lineages: West African, East/Central/South African (ECSA), and Asian. Adaptation to *Ae. albopictus* is strongly lineage dependent. Envelope mutations such as E1-A226V can increase transmission fitness in some genetic backgrounds, whereas epistatic constraints can limit this phenotype in others^2^. Globally, CHIKV causes an estimated 35 million infections annually, with nearly 2.8 billion people living in areas at risk of infection^3^. Climate change, urbanization, and human mobility are further expanding the ecological and geographic conditions suitable for CHIKV transmission^4,5^.

China is considered non-endemic for CHIKV and has historically reported mainly travel-associated infections, with only occasional local outbreaks, including in Guangdong in 2010^6^ and Yunnan in 2019^7^. In early July 2025, however, local transmission was detected in Foshan, Guangdong Province, after which the outbreak subsequently spread across Guangdong and into other provinces of China. More than 25,000 confirmed cases were reported, although the true burden was likely substantially higher as a serological survey in Guangzhou (the capital city of Guangdong Province with a population of 19 million) reported an attack rate of 22%^8^. The 2025 Guangdong CHIKV outbreak thus represents one of the largest documented CHIKV epidemics in a setting previously considered outside the virus’s established endemic range.

Early genomic investigations indicated that the 2025 Guangdong outbreak was caused by an ECSA-lineage virus related to strains from the recent resurgence on Réunion Island^9–12^. CHIKV genomes were also recovered from *Ae. albopictus* in Guangdong, while *Ae. Aegypti* was not detected in entomological surveys^13^, consistent with the likely role of *Ae. albopictus* in local transmission^14^. Preliminary phylogeographic analyses suggested an African origin for the outbreak lineage^15^. Together, these early findings suggested that the virus might have originated from Réunion Island. However, despite these early insights, the timing of introduction, extent of pre-detection transmission, and spatial dynamics of CHIKV establishment in Guangdong remain incompletely resolved, largely because previous analyses were based on limited whole-genome sampling from the outbreak.

Here, as part of the public health investigation of the 2025 Guangdong outbreak, we generated more than 1,200 complete or near-complete CHIKV genomes and analyzed them together with epidemiological, entomological, and mobility data. This integrated genomic epidemiological approach allowed us to reconstruct the likely source and route of viral introduction, estimate the timing of the origin of the sampled outbreak lineage and the period preceding outbreak recognition, and resolve the heterogeneous spatial spread of CHIKV across Guangdong. Our results show how genomic surveillance and phylodynamic inference can reveal otherwise hidden phases of epidemic establishment, and underscore their value as part of integrated early-warning systems in regions at the expanding margins of arbovirus transmission^16^.

## Results

### Chikungunya outbreak in Guangdong, China

Guangdong Province experienced a local chikungunya outbreak in 2010, caused by the ECSA-Indian Ocean Lineage (IOL)^6^, which sits within the broader ECSA lineage diversity, but no large-scale autochthonous transmission was recorded in the subsequent 14 years. On 8 July 2025, the first CHIKV reported case was detected in Foshan, after which cases increased rapidly and autochthonous transmission was reported from all 21 prefecture-level cities within two months (**Fig. 1a, c**). By the end of 2025, 25,584 laboratory-confirmed CHIKV infections had been reported in Guangdong. The epidemic curve comprised two main waves. The first occurred in the summer and was dominated by cases from Foshan (n = 10,853), whereas the second occurred in autumn and was driven primarily by cases from Jiangmen (n = 10,375) (**Fig. 1a**). Following the detection of each wave, vector control measures were implemented in Foshan in early July and Jiangmen in late August, respectively, followed by substantial declines in mosquito ovitrap index (MOI, number of positive ovitraps per 100 retrieved traps) relative to historical baselines (**Fig. 1f-g**).

**Figure 1.**
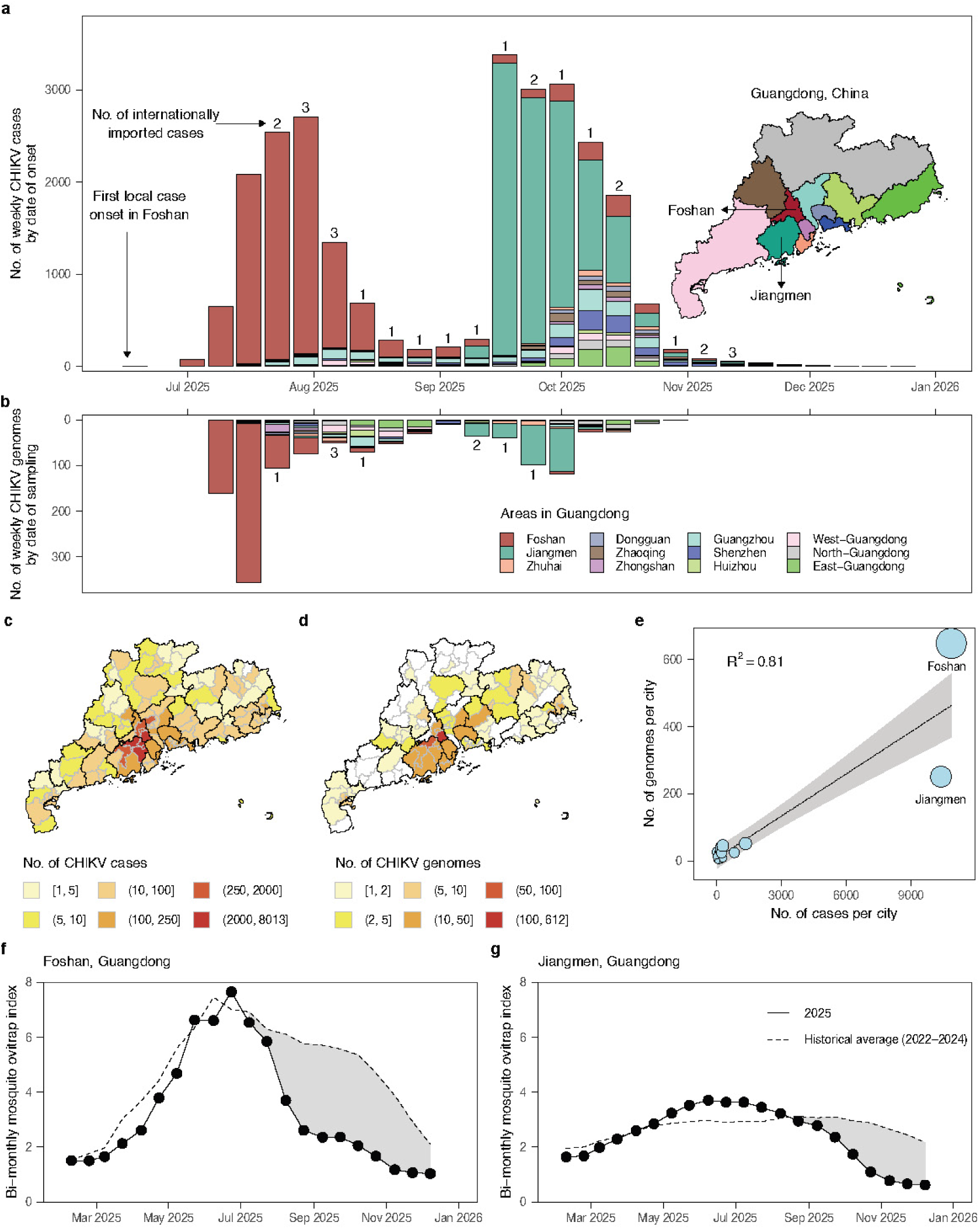
Spatiotemporal distribution of chikungunya virus cases and genomes. (**a**) Epidemic curve of chikungunya virus confirmed cases by date of onset, colored by geographic areas. The number of internationally imported cases are displayed on the bar chart. The inset map shows the administrative boundaries of Guangdong Province, with cities in the western, northern, and eastern parts merged separately. (**b**) Temporal distribution of chikungunya virus genomes generated in this study by sampling date. The number of genomes from internationally imported cases are also displayed on the bar chart. (**c**) Spatial distribution of chikungunya virus cases at the district level. (**d**) Spatial distribution of chikungunya virus genomes at the district level. (**e**) Correlation between the number of CHIKV confirmed cases against the number of CHIKV genomes generated in this study for each of the 21 prefecture-level cities in Guangdong. Circle sizes are proportional to the number of genomes. (**f-g**) Bi-monthly mosquito ovitrap index (MOI) in 2025 compared with the 2022–2024 historical averages for Foshan (**f**) and Jiangmen (**g**), Guangdong. The lines are presented as central-aligned rolling averages. The gray shaded areas represent the reduction in mosquito density in 2025 potentially due to vector control measures.

### Sample collection and genome sequencing

RT-qPCR screening of serum samples from patients attending local designated hospitals identified 7,852 positive samples. We selected positive samples based on cycle threshold (C_t_) values for amplicon-based sequencing using a newly developed pan-lineage CHIKV primer set^17^. This generated 1,264 complete or near-complete consensus genomes (**Extended Data Fig. 1**), with genome coverage ranging from 76.1% to 99.9% (**Fig. S1** and **Fig. 2b**). The median interval between symptom onset and sample collection was 1 day (**Extended Data Fig. 2b**).

**Figure 2.**
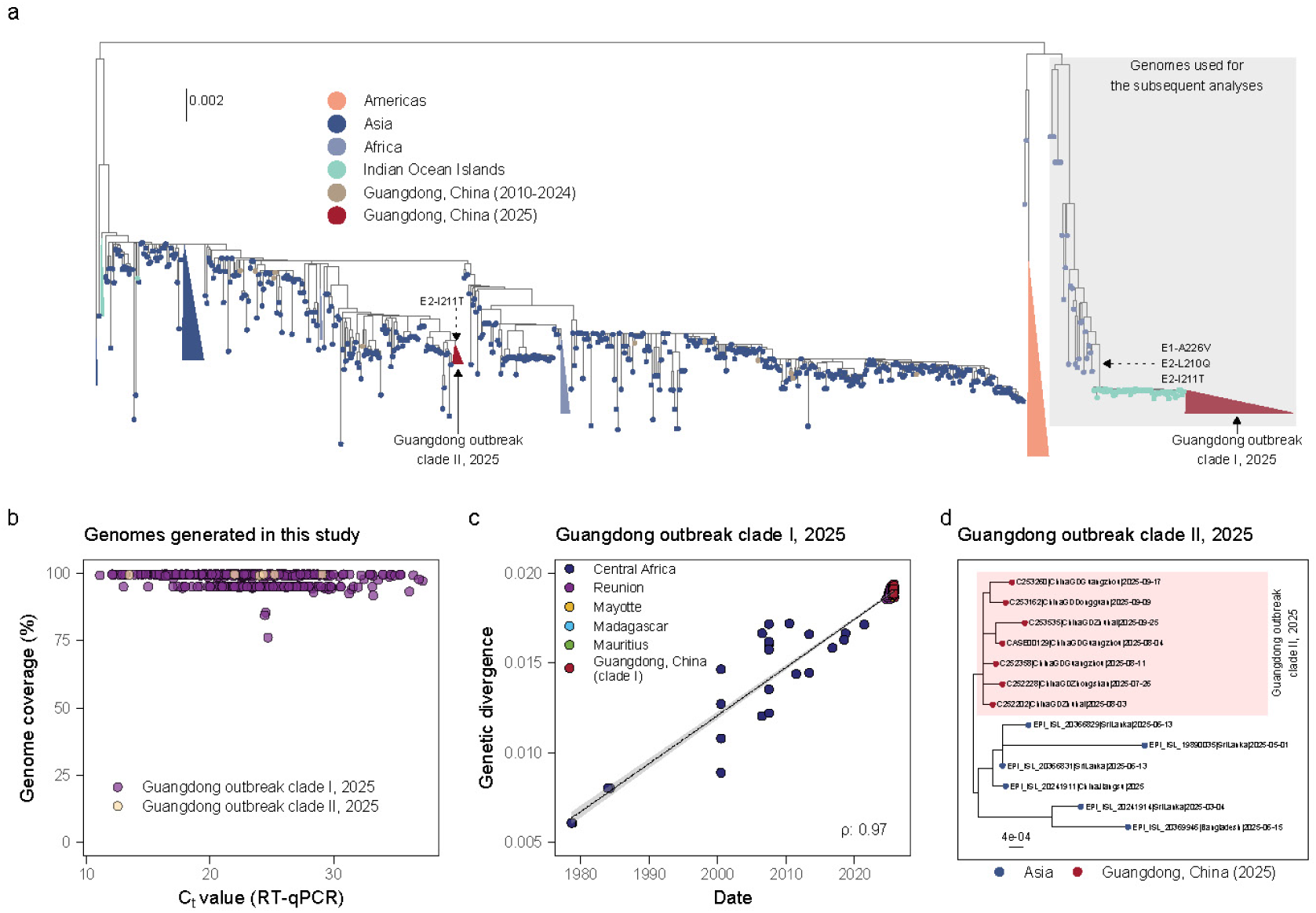
Molecular phylogenetics of the Guangdong chikungunya outbreak in 2025. (**a**) Maximum likelihood phylogeny of chikungunya virus genomes showing two outbreak clades (clade I and clade II; red triangles) during the 2025 chikungunya outbreak in Guangdong. For clarity, some background tips from the same regions are also collapsed as triangles. The scale bar is in units of substitutions per site. (**b**) The genome coverage of chikungunya virus plotted against RT-qPCR C_t_ value. Each point refers to a genome sequenced from an infected case and is colored by the clade type. (**c**) Root-to-tip regression of sampling date of the outbreak clade I genomes alongside the phylogenetically close background genomic sequences (colored according to the sampling region) against genetic divergence, with the Pearson correlation coefficient reported. (**d**) The zoomed view of Guangdong outbreak clade II showing its phylogenetic association with sequences from South Asia.

The spatiotemporal distribution of the newly generated genomes closely mirrored that of reported CHIKV cases (R^2^: 0.81; **Fig. 1e**), capturing distinct epidemic peaks in Foshan and Jiangmen (**Fig. 1b**). Sequencing was concentrated during the early phase of the outbreak, resulting in relative oversampling of Foshan relative to the later Jiangmen wave (**Fig. 1b**; **Fig. 1e**). Nevertheless, the age- and sex-distribution of sequenced cases closely matched those of all confirmed cases (**Fig. S2**), indicating that the genomic dataset broadly represented the demographic structure of the outbreak.

### Phylogenetic placement of outbreak genomes

To place the Guangdong outbreak in a broader genomic context, we combined the newly generated genomes with publicly available CHIKV genomes, including sequences from contemporaneous outbreaks elsewhere and historical genomes from Guangdong. We also generated three additional historical Guangdong genomes, allowing us to assess whether the 2025 outbreak descended from previously detected local diversity.

Nearly all newly generated outbreak genomes were identified as ECSA-2 lineage viruses (99.9%, 1,263/1,264), whereas the remaining genome belonged to the Asian lineage. Focusing on the Guangdong ECSA-2 outbreak, we inferred a maximum likelihood phylogeny from 2,600 CHIKV ECSA genomes (**Fig. 2a**), comprising 1,263 generated during the outbreak, three historical Guangdong genomes sampled before 2025, and 1,334 publicly available genomes. We then defined a Guangdong 2025 outbreak dataset of 1,291 genomes by combining the 1,263 newly generated ECSA-2 genomes with 28 publicly available Guangdong genomes sampled in 2025. Of these, 1,284 genomes, representing 99.5% of the Guangdong outbreak dataset, formed a single monophyletic cluster, hereafter referred to as clade I. The dominant Guangdong outbreak clade, clade I, was most closely related to CHIKV ECSA genomes from the Indian Ocean region, particularly Réunion^18^ (**Fig. 2a**). Notably, all clade I genomes encoded the E1-A226V substitution, a well-characterized determinant of increased CHIKV transmission by *Ae. albopictus*. This mutation was not acquired after arrival in Guangdong but was already present in the 2024–2025 Réunion epidemic lineage, together with other substitutions linked to enhanced *Ae. albopictus* fitness, including E2-I211T and E2-L210Q^19,20^ (**Fig. 2a**).

The remaining seven Guangdong genomes fell outside clade I and instead formed a distinct cluster, clade II (**Fig. 2a**). Within this clade, the Guangdong genomes were most closely related to sequences from South Asia, including Sri Lanka and Bangladesh (**Fig. 2d**). This phylogenetic placement was consistent with epidemiological investigations, as all seven clade II cases were classified as imported and had recent travel histories to Bangladesh. Although these introductions occurred after mid-July, clade II did not appear to have established sustained onward transmission or displace the locally established clade I lineage (**Extended Data Fig. 3**). Notably, clade II genomes lacked the E1-A226V and E2-L210Q substitutions, which may have contributed to their failure to establish sustained onward transmission. Overall, our findings indicate that, despite multiple introductions of CHIKV into Guangdong, the epidemic was driven overwhelmingly by a single successful introduction of a Réunion-associated, *Ae. albopictus*-adapted ECSA lineage.

### Source and route of the dominant outbreak lineage

The clade I dataset with phylogenetically close background genomic sequences showed strong temporal structure (**Fig. 2c**; ρ = 0.97). We therefore used a Bayesian phylogeographic model to reconstruct the time-scaled phylogenetic history of the dominant Guangdong outbreak lineage and infer ancestral locations along the phylogeny (**Fig. 3**). The resulting maximum clade credibility tree was largely consistent with the ML phylogeny (**Fig. 2a**) and previous studies^9,10,15^, and placed all randomly sub-sampled Guangdong sequences in a single well-supported clade (posterior probability = 1.00) (**Fig. 3a**). Discrete geographic reconstruction identified Réunion as the most likely location of the common ancestor of clade I (**Fig. 3a**; location posterior probability = 1.00), which aligns with the previous findings^9–12^. The same analysis also identified Réunion as a regional hub for onward spread to other Indian Ocean islands, including Madagascar, Mauritius and Mayotte (**Fig. 3a**; robust to the down-sampled numbers of Réunion genomes: **Extended Data Fig. 4**). More broadly, our analysis was consistent with long-term circulation of the ancestral lineage in Central Africa, followed by its emergence in Réunion after mid-2024, and subsequent introduction into southern China through Foshan (**Fig. 3a**, inset map). From this inferred point of entry, the virus spread across Guangdong Province (**Fig. 3b**).

**Figure 3.**
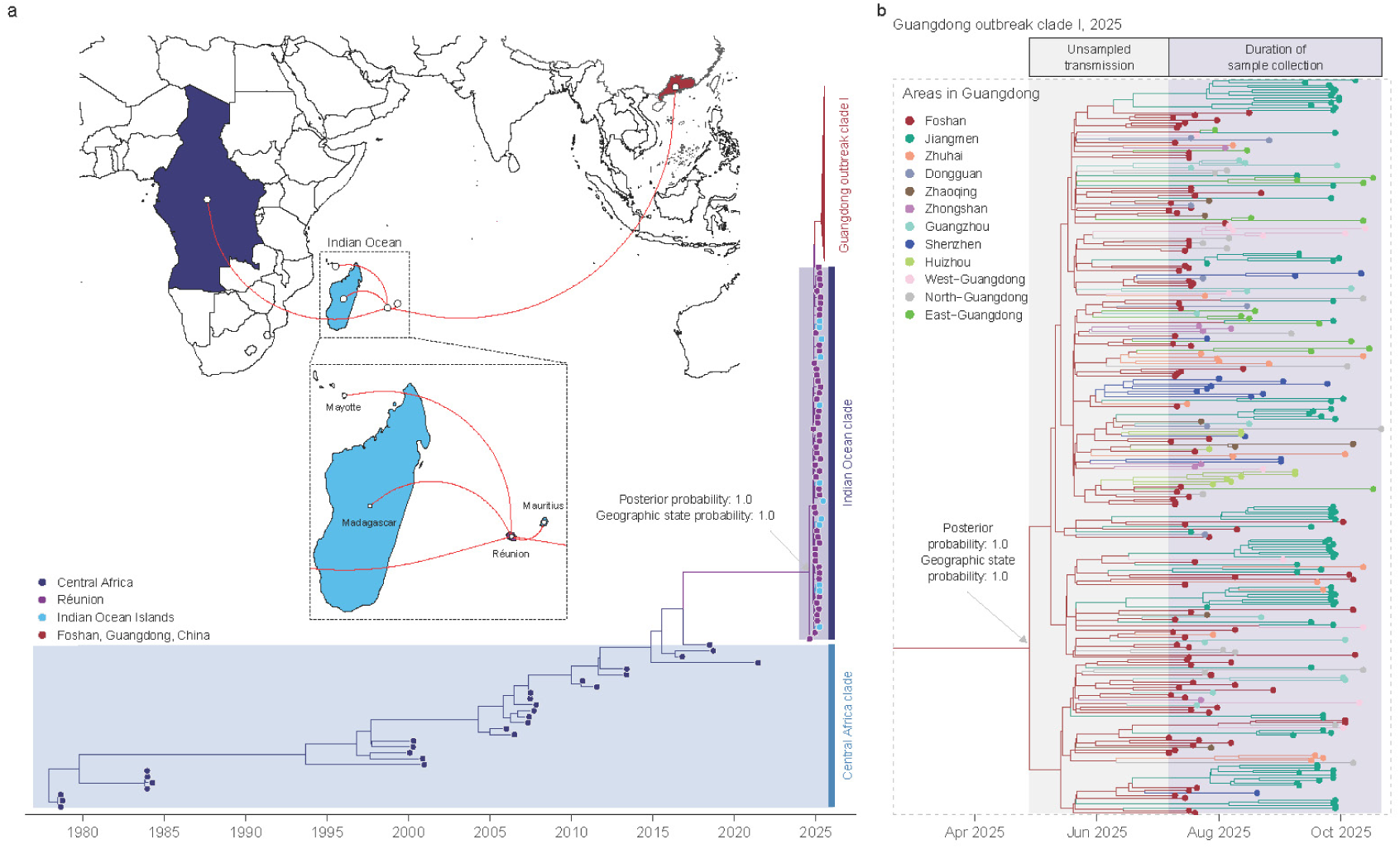
Genomic epidemiology reconstructs the introduction of chikungunya virus into Guangdong, China. (**a**) Maximum clade credibility phylogeny inferred under a discrete phylogeographic model. Tips are colored according to the sample locations and branches are colored by the inferred geographic states. The posterior probabilities and geographic state probabilities for selected nodes are labelled. The inset map shows the inferred dispersal patterns from Central Africa, across the Indian Ocean, to Guangdong, China. (**b**) Detailed view of the Guangdong outbreak clade I.

### Timing of introduction and pre-detection transmission

To estimate the timing of the origin of the sampled Guangdong outbreak lineage, we inferred the time of the most recent common ancestor (tMRCA) of clade I. In the main phylogeographic analysis, the clade I tMRCA was estimated as 28 April 2025 (95% highest posterior density [HPD] = 20 April 2025 to 12 May 2025) (**Fig. 4a**), while its parent node was dated to 27 December 2024 (95% HPD = 31 October 2024 to 11 April 2025). The introduction into Guangdong is therefore likely to have occurred along the ancestral branch leading to clade I. The clade I tMRCA represents the latest plausible bound for this introduction.

**Figure 4.**
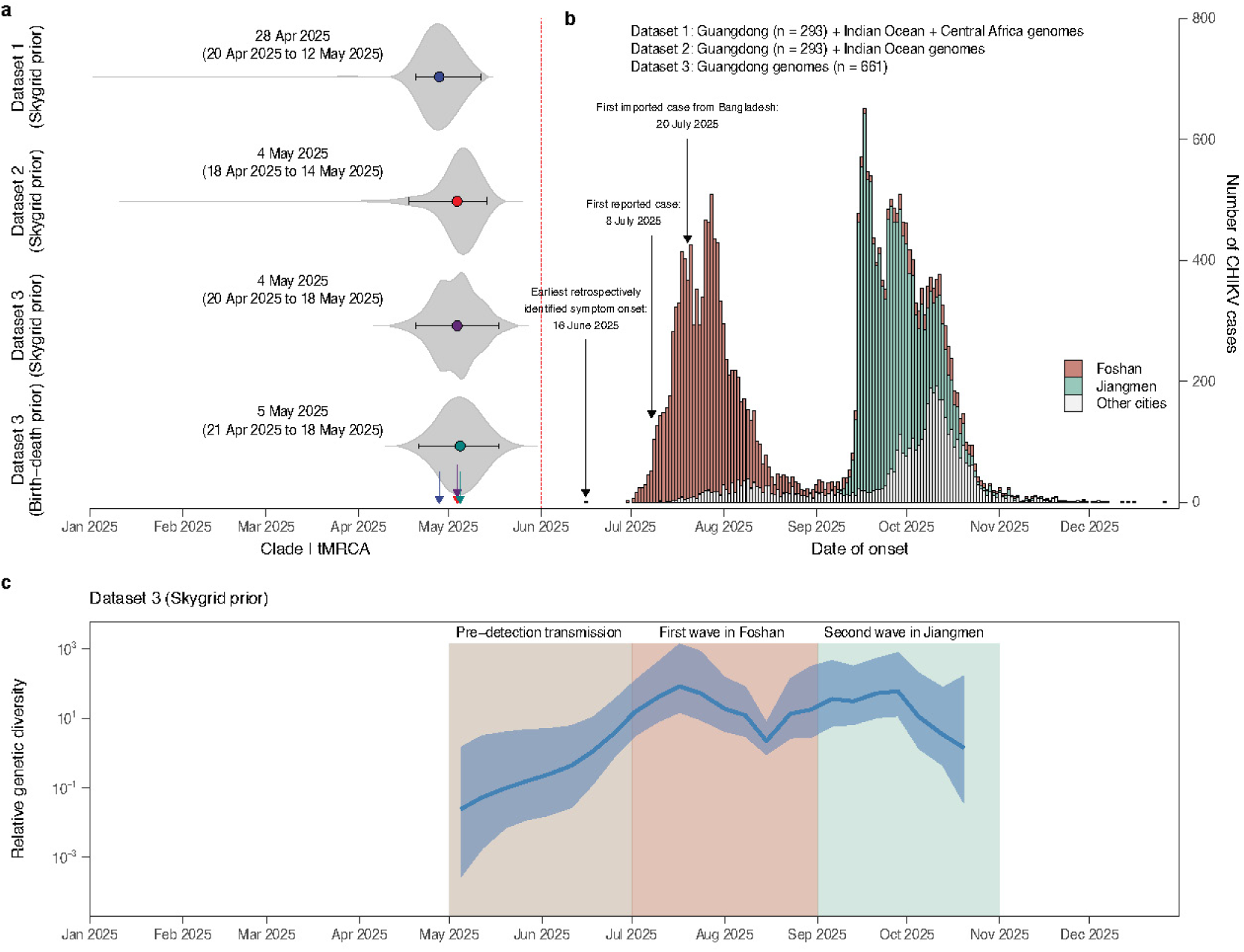
The time of the most recent common ancestor (tMRCA) of the outbreak clade. **I.** (**a**) Violin plots showing estimated posterior distributions of the tMRCA of the outbreak clade I. Points and ranges represent the posterior mean and 95% HPD intervals, respectively. Estimates were obtained using three different datasets (datasets 1-3, with details in panel **b**) and under different tree prior models. (**b**) Epidemic curve of chikungunya virus cases by date of onset. (**c**) Relative genetic diversity of chikungunya viruses, as inferred by Bayesian Skygrid population analysis of dataset 3 (n = 661 outbreak genomes).

The estimated tMRCA of clade I was robust to the genomic dataset, nucleotide substitution model, molecular clock model and tree prior, with posterior means ranging from 27 April to 5 May 2025 and overlapping 95% HPD intervals across all configurations (**Fig. 4a, Table S2**). Analyses containing background genomes dated the parent node of clade I to late December 2024, although the broad 95% HPD intervals extended from October 2024 to April 2025. Furthermore, this inferred introduction window overlapped with the Spring Canton Fair, held in Guangdong from 15 April to 5 May, a major international event associated with substantial international travel (**Fig. 5c**). Although the earliest confirmed case was reported on 8 July, and the earliest retrospectively identified symptom onset was traced to 16 June, our analyses place the origin of the outbreak clade I approximately 1.5–2.5 months earlier (**Fig. 4a-b**). Clade I may therefore have circulated locally for several weeks before detection. Imported cases were not detected until 20 July, further underscoring the limited sensitivity of passive surveillance to detect the early establishment of clade I in Guangdong.

**Figure 5.**
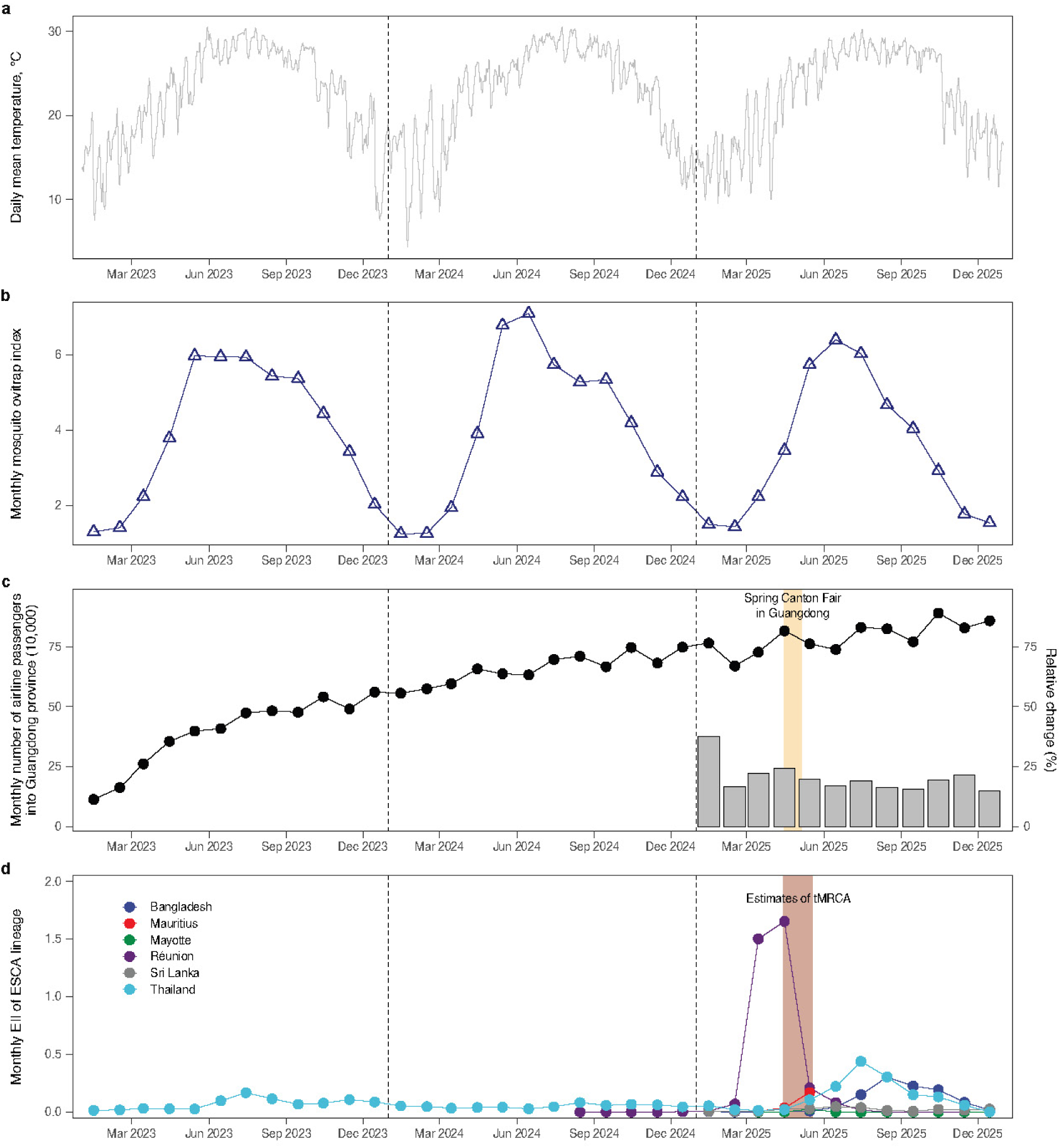
Time series of potential factors driving the emergence of chikungunya outbreak in Guangdong, China. **(a)** Temporal dynamic of daily mean temperature in Guangdong, which was retrieved from ERA5^21^. (**b**) Mosquito ovitrap index (MOI, number of positive ovitraps per 100 retrieved traps) averaged across Guangdong. (**c**) Global number of airline passengers into Guangdong Province. Gray bars denote the monthly change of airline passengers in 2025 relative to that in 2024. Data of air passenger volumes were provided by Official Airline Guide (OAG) Ltd. (**d**) Estimated importation intensity (EII) from selected countries in Africa and Asia. The light-red shaded area represents the combined 95% HPD intervals of clade I tMRCA across four independent analyses.

Reconstruction of viral population dynamics further revealed that the establishment of clade I was followed by a gradual increase in relative genetic diversity from early May, before the first reported case. Relative genetic diversity peaked twice, in mid-July and late-September (**Fig. 4c**), closely following the Foshan and Jiangmen epidemic waves. We therefore refer to May–June 2025 as a putative phase of pre-detection transmission, preceding the two recognized epidemic waves (**Fig. 4c**).

### Mobility context and importation intensity

To assess whether independent ecological and mobility data were consistent with the timing of introduction from Réunion, we examined local temperature, mosquito abundance and international air traffic volumes into Guangdong from 2023 to 2025 (**Fig. 5a-c**). Temperature and mosquito abundance were broadly comparable to those observed in previous years. In contrast, international passenger arrivals into Guangdong increased during April and May 2025, coinciding with the Spring Canton Fair (**Fig. 5c**). These observations suggest that the inferred date of introduction was characterized not only by local climatic and entomological conditions, but with increased opportunities for viral importation during concurrent CHIKV activity abroad.

We thus calculated the estimated importation intensity (EII) of the CHIKV ECSA lineage into Guangdong across source locations with available data (**Fig. 5d**). This metric combined CHIKV incidence at source locations, the relative frequency of ECSA-lineage genomes and monthly air passenger volumes into Guangdong, yielding a relative measure of importation intensity. Réunion showed a sharp peak in EII in April 2025 that was primarily driven by a surge of local case prevalence (**Fig. 5d, Fig. S3**), further supporting the introduction window inferred from genomic dating and phylogeographic analysis.

### Spatial expansion and heterogeneous onward diffusion

Following its introduction, clade I established local transmission chains that were initially concentrated in Foshan and neighboring cities. Discrete phylogeographic reconstruction showed that viral movement from Foshan to Jiangmen peaked in late June and gradually decreased through July (**Fig. 6a**). In contrast, viral exports from Jiangmen to other cities were limited during the second wave, suggesting that the Jiangmen outbreak was driven mainly by local amplification rather than extensive onward dissemination. We identified statistically supported viral dispersal routes (Bayes factor >3), including multiple routes originating in Foshan and a single route from Jiangmen to Zhuhai (**Fig. 6b**). Viral movement out of Foshan was already evident during the pre-detection transmission phase (**Fig. 6c**), indicating that spatial dissemination preceded the first case notification.

**Figure 6.**
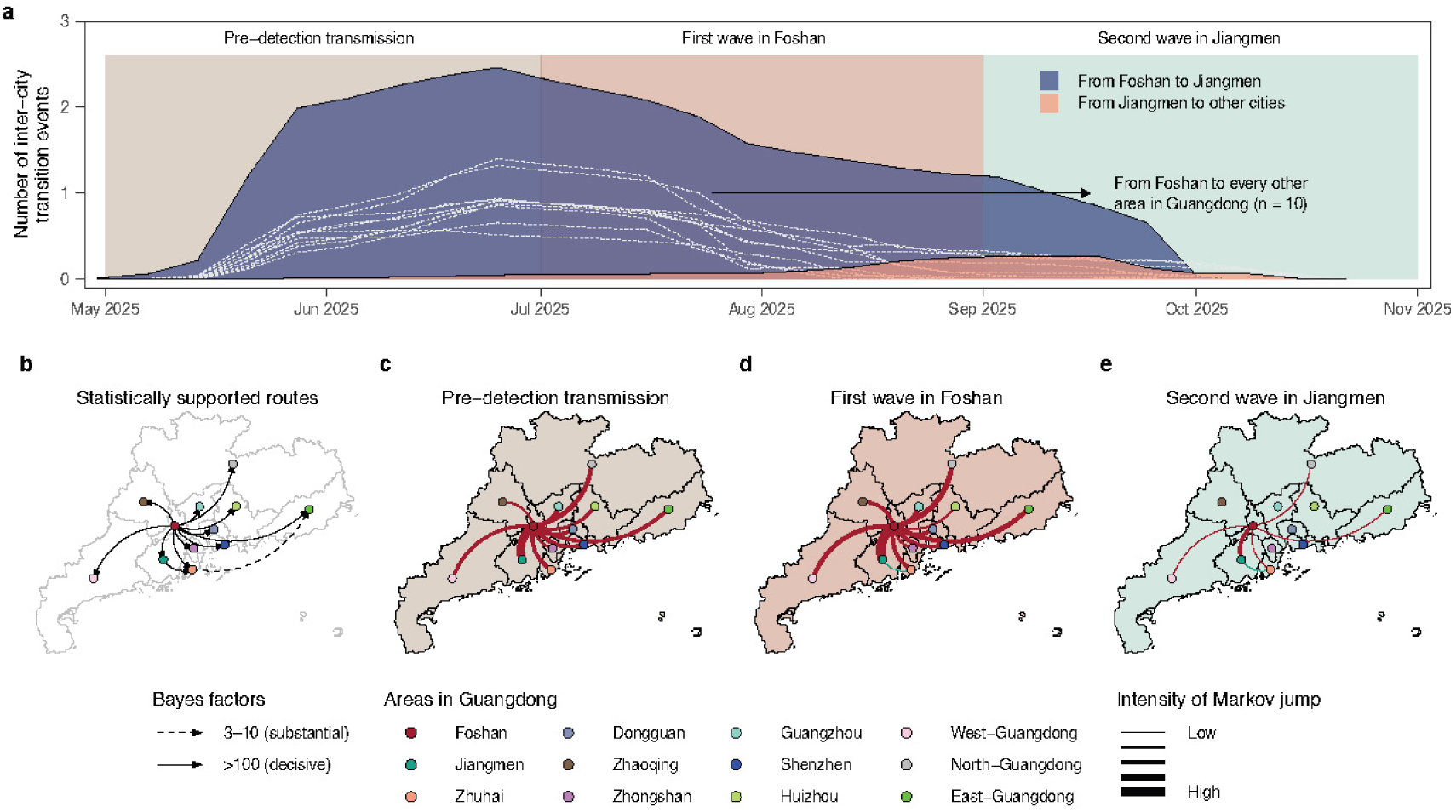
Discrete phylogeographic reconstruction of inter-city CHIKV movement during the Guangdong outbreak, China. (**a**) Weekly inter-city Markov jump counts (location transition events) through time. (**b**) Migration routes with statistically supported evidence. (**c-e**) Estimated number of location transition events per period between each pair of geographic areas. Line thickness denotes the intensity of Markov jumps.

To resolve local spatiotemporal dynamics, we also performed spatially explicit phylogeographic reconstructions using a prevalence-adjusted subsample of 661 clade I genomes. Following sub-sampling, the spatiotemporal distribution of genomes was representative of reported cases across Guangdong (R^2^: 0.99; **Extended Data Fig. 5**). This analysis revealed a multiscale expansion process combining short-distance local spread with longer-distance dispersal across Guangdong. In May 2025, inferred viral dispersal was restricted to Foshan, with all movements occurring within the city (**Figs. 7a**, **7f**). By June, clade I had spread to neighboring urban conurbations, including Guangzhou and Zhongshan (**Fig. 7b**). Inter-city viral movement gradually became dominant by late July, when the proportion of within-city movements declined to 34.5%, compared with more than 90% during May–June 2025 (**Fig. 7f**). This temporal shift was consistent with a transition from localized early transmission to broader, human-mediated dissemination across provincial transportation networks (**Fig. 7a**).

**Figure 7.**
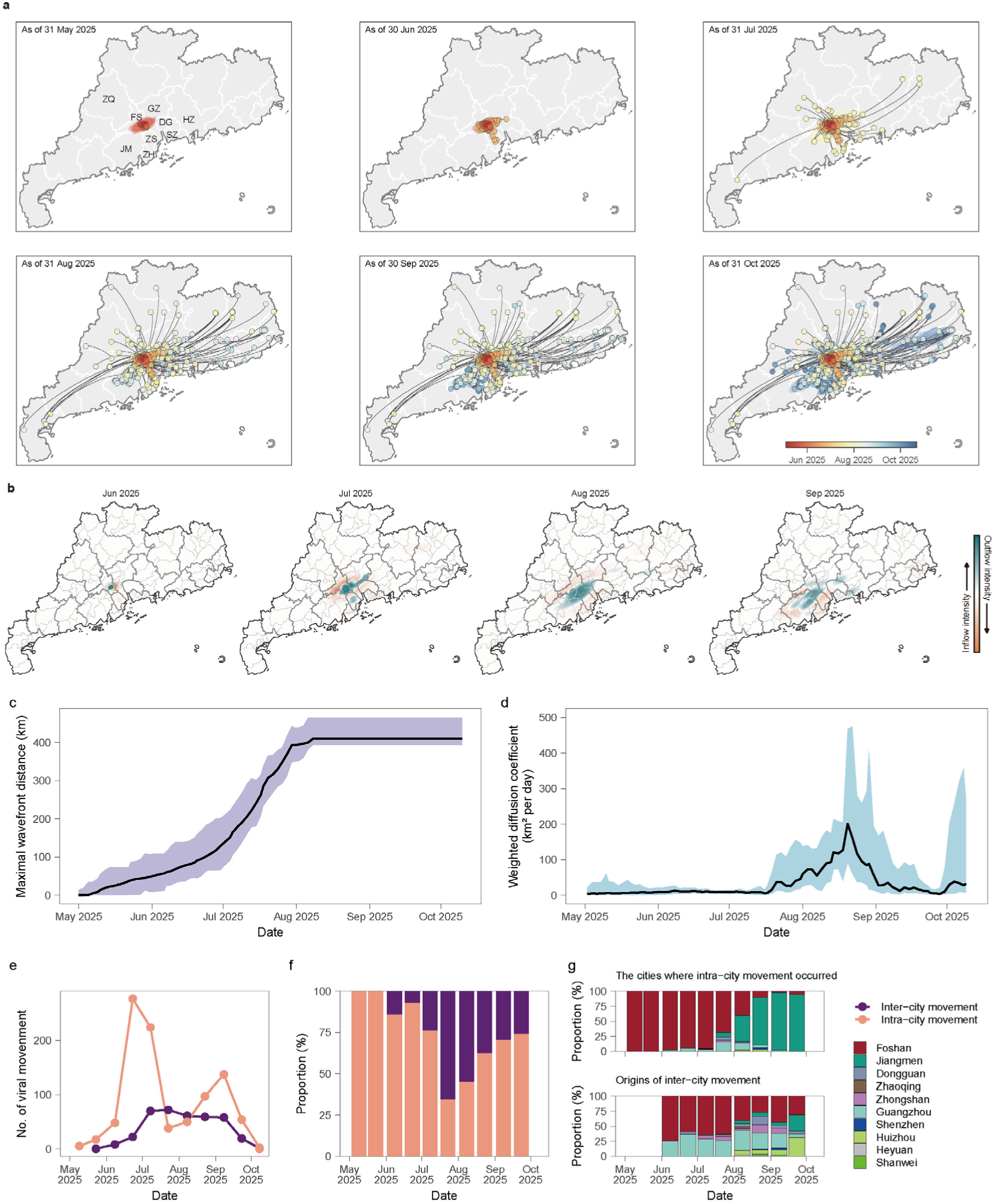
Continuous phylogeographic reconstruction of CHIKV spatiotemporal diffusion across Guangdong, China. (**a**) Maps display the spatially explicit dispersal history of the outbreak lineages until various time points. Points represent nodes of the maximum clade credibility (MCC) tree. The 80% highest posterior density (HPD) regions behind the points reflect the inference uncertainty summarized from 1,000 posterior tree distributions and were computed for successive time layers. Nodes and uncertainty regions are colored according to time. Curves represent dispersal events with the direction of counter clockwise from origin to destination. City codes: FS, Foshan; JM, Jiangmen; ZH, Zhuhai; DG, Dongguan; ZQ, Zhaoqing; ZS, Zhongshan; GZ, Guangzhou; SZ, Shenzhen; HZ, Huizhou. (**b**) The spatial inflow and outflow hotspot of outbreak lineages fitted from the Gaussian kernel density estimation (KDE). Blue and red colors refer to areas with high intensity of viral outflow and inflow, respectively. (**c**) Median maximal wavefront distance (km) of outbreak lineages over time. The shaded areas denote the 95% HPD. (**d**) Median weighted diffusion coefficient (km^2^ per day) of outbreak lineages over time. (**e-f**) The inter-city (purple line) and intra-city (orange-pink line) movement intensity from May to October 2025. (**g**) The top panel denotes the proportion of cities where intra-city movement occurred, and the bottom panel refers to the proportion of geographic origins of inter-city movement events.

However, longer-distance seeding did not result in sustained transmission in all recipient locations. Jiangmen was the main exception, becoming a second major transmission focus in August and September. Unlike Foshan, transmission in Jiangmen was characterized predominantly by sustained short distance spread, indicating a more spatially concentrated second wave (**Fig. 7b**). This difference may reflect the timing and intensity of control measures, local vector ecology (a relatively lower mosquito ovitrap index in Jiangmen; **Fig.1 f-g)**, population movement patterns or heterogeneity in population susceptibility.

We then quantified the extent of spatial dissemination of clade I across Guangdong. The maximal wavefront distance, defined as the greatest distance reached by phylogenetic branches from the inferred epidemic origin, increased steadily between May and July 2025 (**Fig. 7c**). By the peak of the first epidemic wave in late July 2025, the outbreak had expanded ∼ 400 km from its epidemic origin, corresponding to an average expansion rate of ∼ 4.3 km/day. The weighted diffusion coefficient increased from mid-July to mid-August (**Fig. 7d**), corresponding to the geographic expansion phase of the Foshan wave, and peaked at 200 km^2^/day (95% HPD: 92 to 468) in mid-August (**Fig. 7d**). No comparable spatial expansion was observed during the Jiangmen wave, further supporting the interpretation that the second wave was more geographically localized.

The two epidemic waves also differed in their movement structure. Peaks in within-city movement occurred during both waves, whereas high intensity inter-city movement was concentrated between July and August (**Fig. 7e**). Foshan therefore acted both as a local amplifier and as a source of longer-distance viral dissemination. In contrast, Jiangmen functioned primarily as a source of within-city spread. Guangzhou, the capital city of Guangdong Province, also contributed substantially to inter-city viral movement, accounting for 25.0-36.4% of inferred inter-city movements during June-August 2025 (**Fig. 7g**). This finding is consistent with Guangzhou’s role as a major provincial transportation hub in Guangdong Province and with epidemiological reports of cross-city travel histories^22^. Overall, these results show that CHIKV spread across Guangdong was highly heterogeneous, with a small number of cities seeding most longer-distance movements, whereas most cities acted primarily as sinks with limited onward export.

## Discussion

Our integrated genomic, epidemiological, and entomological investigation reconstructs the establishment and spatiotemporal expansion of the largest documented CHIKV outbreak in China to date. The outbreak marks an important shift from the sporadic, travel-associated infections that have so far characterized CHIKV in southern China to explosive, sustained transmission across multiple cities. We show that this epidemic was driven predominantly by a single successful introduction of an *Aedes*-*albopictus* adapted lineage rather than repeated introductions. Our genomic reconstructions support a period of several weeks of undetected transmission before the first case was reported. By resolving the timing and drivers of introduction alongside the heterogeneity of onward spread, our analysis extends earlier genomic reports of the outbreak^9–11^ and shows how phylodynamic inference can recover phases of the epidemic establishment that are not detectable using case-based surveillance.

Guangdong received at least two CHIKV introductions during this period, clade I from Réunion and clade II from South Asia. Yet only clade I established sustained transmission, in contrast to the multiple co-circulating introductions that seeded the contemporaneous Mauritius outbreak^23^. Clade I carried the *Ae. albopictus* adaptive substitutions E1-A226V and E2-L210Q and arrived in Foshan during a window of high human mobility and vector activity, whereas the clade II importations lacked these substitutions and arrived under intensified case management and vector control, resulting in no onward transmission. Whether a single introduction ignites a local epidemic depends on a complex interplay of viral genotype, competent vector abundance, climate, and population susceptibility. Our earlier semi-mechanistic analysis of the 2014 Guangzhou dengue outbreak showed that establishment required the simultaneous alignment of imported cases, mosquito density, and temperature, with a deficit in any singular factor being sufficient to prevent it^24^. Likewise, a recent analysis of 86 CHIKV outbreaks attributed much of the variance in outbreak size and timing to stochasticity^25^. Disentangling these determinants is increasingly important given growing evidence of lineage-specific differences in transmissibility and clinical presentation, and will require integrated epidemiological, ecological, and virological data.

In Guangdong Province, active surveillance of dengue virus is conducted annually from April to November, whereas early detection of chikungunya virus mainly relies on the passive surveillance system. The interval between the estimated origin of the sampled outbreak lineage and the first reported case highlights a structural limitation of clinician-initiated passive surveillance in regions at the margins of endemic transmission. Larger windows of one year have been reported during the emergence of the Zika virus epidemic in Brazil^26^. Although the ECSA lineage has a relatively low inapparent fraction (∼18%, compared with ∼48% for the Asian lineage^27^), clinicians in a non-endemic setting are unlikely to suspect CHIKV, particularly because early cases may be mild and their presentation overlaps with co-circulating viruses. Multi-pathogen arbovirus sentinel surveillance could provide an important complement, which captures more detailed clinical data^28^ and has been shown to precede reporting in passive surveillance systems by up to three and eight weeks for dengue and Zika, respectively^29^. Adding genomic surveillance to sentinel systems would extend their reach from detecting cases earlier to identifying which lineages are circulating and how long transmission has gone undetected.

Our findings of viral introduction and pre-detection transmission relied on viral genome-based phylodynamic inference, an approach that has been applied to Zika^30^ and yellow fever epidemics^31^. Although direct evidence of cryptic transmission is not yet available, several independent observations support this interpretation. First, the estimated timing of the clade I tMRCA was highly consistent across genomic datasets, substitution models, molecular clock models and tree priors (**Table S2**). Second, the inferred tMRCA coincided with a peak in estimated importation intensity from Réunion (**Fig. 5d**) and with the seasonal increase in mosquito abundance in Guangdong (**Fig. 5b**). Third, relative genetic diversity began increasing before the first reported case was identified (**Fig. 4c**), consistent with local transmission preceding outbreak recognition. Nevertheless, direct confirmation of transmission before June 2025 would require retrospective testing of clinical or surveillance specimens collected during that period. These findings further demonstrate the value of expanding active genomic surveillance in regions at the fringe of CHIKV endemic transmission, where earlier detection of transmission could improve outbreak preparedness and response.

Our study also showed that the CHIKV outbreak in Guangdong was characterized by a highly heterogeneous spatial expansion, with direct implications for control. The two epidemic waves differed fundamentally in structure. The Foshan-centred first wave combined local amplification with medium- and long-range inter-city dispersal along provincial transport networks, whereas the Jiangmen-centred second wave was sustained largely by short-distance local spread. Previous epidemiological analyses also showed varying epidemic half-lives between the two waves in Guangdong^32^. A small number of cities, notably Foshan and the transport hub Guangzhou, seeded most longer-range movements, while the majority acted as transmission sinks with limited onward export. This source–sink structure suggests that interventions concentrated on a few well-connected hub cities early in an outbreak could disproportionately limit province-wide spread, and that the appropriate control strategy differs between an inter-city dissemination phase and a locally amplifying wave.

These dynamics are not unique to Guangdong. Under changing climate and expanding vector suitability, CHIKV is re-emerging globally across the Americas^33,34^, Africa^18,23^, Europe^4^, South-east Asia^35^, and southern China^36^. Situational awareness must shift from a single-country view to tracking geographic expansion and the transition of receptive, competent-vector regions toward endemicity. Yet, global CHIKV surveillance remains less coordinated than for dengue, for which platforms such as OpenDengue^37^ and the WHO Global Dengue Dashboard^38^ have been established to enhance preparedness and monitoring of epidemic trends. We highly recommend integrating global CHIKV surveillance into the existing flavivirus surveillance network. A comparable framework for CHIKV, coupled with routine genomic data sharing, would enable real-time tracking of geographic spread and vector-adaptive evolution, supporting timely alerts to emerging outbreaks.

Our phylodynamic results should be interpreted in light of several limitations. First, despite our prevalence-informed subsampling, the spatial and temporal distribution of genomes was not perfectly representative, reflecting heterogeneity in sequencing intensity across cities. The clade I tMRCA represents the latest plausible bound for introduction, rather than the precise date on which CHIKV entered Guangdong, and should be interpreted together with its 95% HPD interval and the estimated date of the parent node. Second, phylogenetic placement relies on publicly available background genomes and may shift as additional genomes from Réunion, the wider Indian Ocean and South Asia become available. Therefore, the role of Réunion in the CHIKV dispersal network should be interpreted within the context of current data availability and sub-sampling scheme. Although our sensitivity analyses showed the robustness of down-sampling genomes from Réunion (**Extended Data Fig. 4**), a comprehensive understanding of Réunion’s role will require more geographically representative genomic sampling from other regions. Third, the mechanisms underlying the twin Foshan and Jiangmen waves remain unresolved and may reflect spatial heterogeneity in local environment, vector abundance, population susceptibility, control measures or reporting. These questions warrant mechanistic models integrating phylogenetics with transmission models at the vector-human interface^39^.

## Conclusion

The 2025 Guangdong outbreak demonstrates how a single viral introduction can establish sustained transmission and generate a large epidemic in a setting previously considered non-endemic for CHIKV. The genomic evidence supporting transmission before outbreak recognition highlights the limitations of passive surveillance at the margins of endemic transmission. As climate, vector distributions, and human mobility reshape the geography of arboviral risk^40^, integrating sentinel, genomic, and mobility-informed surveillance will be central to detecting the transition from importation to establishment, a priority now recognized in global frameworks for arbovirus epidemic and pandemic preparedness^41^ and increasingly urgent across the growing number of regions receptive to arboviral emergence.

## Methods

The institutional review board at Guangdong Provincial Center for Disease Control and Prevention waived ethical approval for this work (Ethical Exemption Number: W96-027M-202502), as it determined that our project is based on residual serum as part of public health surveillance and all analyzed data were anonymized.

### Chikungunya virus case data

The date of first symptom onset in the 2025 Guangdong chikungunya virus outbreak was recorded as 16 June 2025, with the first case reporting on 8 July^36^. Surveillance was initiated after detection of the first reported case. International travel-associated (referred to as internationally imported cases hereafter) and locally acquired chikungunya virus cases (referred to as local cases) diagnosed in Guangdong Province after 16 June were extracted from the National Notifiable Infectious Disease Reporting System (NNDRS). For each laboratory-confirmed case, we retrieved the information about sex, age, address of residence, date of symptom onset, and date of reporting where available. Cases with a travel history to Macau and diagnosed in Guangdong were believed to have their exposures locally as the contemporaneous chikungunya transmission in Macau remained at a low level (only 8 local cases were reported in 2025; https://ssm.gov.mo/portal/), thereby classifying them as local cases.

### Entomological surveillance

Surveillance of *Ae. aegypti* and *Ae. albopictus* was conducted in Guangdong on a monthly or bi-monthly basis as part of the national entomological surveillance program, from which the mosquito ovitrap index (MOI) was estimated. MOI was defined as the number of positive ovitraps per 100 retrieved traps.

### Sample collection

We collected serum samples from patients who exhibited symptoms consistent with CHIKV infection at local designated hospitals in Guangdong, China. All specimens were residual aliquots from routine clinical tests. Demographic and epidemiological data, including age, gender, date of symptom onset, and date of specimen collection, were recorded for each patient where available. For specimens with unknown date of specimen collection, we inferred it using the time lag between symptom onset and specimen collection (**Extended Data Fig. 2b**), or between specimen collection and receipt. All serum specimens were immediately stored at -80 °C after collection and maintained under ultra-low temperature until nucleic acid extraction and downstream experiments.

### Nucleic acid isolation and RT-qPCR

Total viral RNA was extracted from serum samples using the commercial nucleic acid detection kits (e.g., Tianlong, Xi’an, China; BioGerm, Shanghai, China; Qingcao Gene, Foshan, China) in accordance with the manufacturer’s standard operating procedures. The purified RNA was used as the template for RT-qPCR to confirm CHIKV positivity. The cycle threshold (C_t_) value of each valid sample was documented to indicate relative viral loads. We collected a total of 7,852 RT-qPCR-positive serum samples from July to December 2025, which were subjected to subsequent whole-genome amplification and high-throughput sequencing. All experiments were conducted in a biosafety laboratory under standard protocols to prevent cross-contamination.

### Amplicon-based sequencing

Positive serum samples with C_t_ values below 30 were primarily selected for sequencing, while a subset of samples with C_t_ values greater than 30 were also sequenced on a case-by-case basis if they were deemed epidemiologically important. The percentage of positive samples sequenced varied over time, with approximately 35% sequenced in July and approximately 10% in September; no sequencing was performed from November 2025 onwards.

First-strand complementary DNA (cDNA) was synthesized from purified viral RNA using the LunaScript RT SuperMix Kit (NEB, Ipswich, MA, USA). Reverse transcription was carried out under the following conditions: 25 °C for 2 min, 50 °C for 10 min, and a final incubation at 95 °C for 1 min to fully inactivate reverse transcriptase. Multiplex long-range PCR was then used to amplify the full-length CHIKV genome with a previously validated pan-lineage primer set described in our previous study^17^. This primer panel contained 10 pairs of overlapping oligonucleotides separated into two independent primer pools, which generated approximately 1200 bp overlapping amplicons spanning the entire ∼11.8 kb CHIKV genome. PCR was performed using Q5 High-Fidelity 2× Master Mix with the thermal profile: initial denaturation at 98 °C for 30 s, followed by 35 cycles of 95 °C for 15 s and 65 °C for 5 min. The resulting PCR amplicons were purified with 0.5× AMPure XP beads (Beckman Coulter, Brea, CA, USA) and eluted in nuclease-free water for library preparation.

Purified amplicons were prepared for library construction using the Nextera XT DNA Library Preparation Kit (96 samples) (Illumina, Cat. No. FC-131-1096) and sequenced on the Illumina MiniSeq platform in either single-end or paired-end mode as required. Raw data in FASTQ format were subjected to quality control using Fastp^42^. The quality-filtered reads were mapped to the reference sequence (GenBank Accession number: NC_004162) using Burrows-Wheeler Alignment Tool and SAMtools^43^. Consensus sequences were generated using iVar v1.4.4^44^, with a minimum sequencing depth threshold of 10× for consensus calling. Positions below the depth threshold were masked with N characters.

Finally, 1,264 complete or near complete consensus genomes were successfully assembled during the outbreak, and we included three additional CHIKV genomes sampled before 2025 in Guangdong, resulting in a total of 1,267 genomes. The heterogeneous genomic surveillance intensity led to the spatially and temporally inconsistent distribution of genomes (**Fig. 1e**), which highlighted the necessity of adopting sub-sampling strategy in the subsequent phylogenetic analyses.

### Global dataset of CHIKV genomes

To contextualize the genetic diversity of the CHIKV outbreak in Guangdong, we combined our complete or near complete genome sequences with a global dataset of publicly available CHIKV genome sequences from GISAID EpiArbo as of 14 April 2026 (https://doi.org/10.55876/gis8.260914ys) ^45^. Public genomes were included in our analysis only after their corresponding publishing embargo had expired. We also retrieved the original entry of CHIKV genomes from the GenBank database to double-check and correct the sampling locations in GISAID due to the possibility of inaccurately recorded location there, especially for those whose travel history was not correctly linked. For publicly available Guangdong genomes generated by other institutions, we paid close attention to genome-related original publications^13,46^, where available, to further correct errors in the entry of location and date of collection in GISAID.

We used the Chikungunya Virus Typing Tool to assign a specific lineage for each CHIKV sequence^47^. Since 99.9% (1,263/1,264) of the Guangdong genomes generated in this outbreak were identified as belonging to the ECSA lineage (the remaining one was assigned as Asian lineage and had a travel history to Thailand), global genomes belonging to West African or Asian lineages were excluded from subsequent analyses. We further discarded sequences lacking metadata on collection date or sampling locations and only retained sequences from urban-cycle hosts. Sequences from multiple passages or from artificially infected *Aedes* were also removed. The sampling location of sequences collected from returning travelers was set to the travel destination where infection likely occurred.

Global background genomes of ECSA lineage were combined with the ECSA genomes generated in this study (n = 1,263), and then aligned to the reference genome (NC_004162) using MAFFT^48^ and inspected manually. Only two open reading frames (ORF) were used and concatenated together after trimming the end regions and the untranslated junction region between the two ORFs. For the global background genomes, we first removed identical post-alignment genomes sampled from the same host type and kept the one with the earliest collection date. Second, only genomes collected from tropical regions (i.e., Africa, America, and Asia) before 2026 were retained. Third, to reduce the number of genomes and account for bias in surveillance intensity across locations and years, we performed a sub-sampling process to assemble the background set. Specifically, a maximum of 20 genomes per country per year were randomly selected from Asia and America, and a maximum of 40 genomes per country per year from Africa, whereas all genomes from China were kept for downstream analyses. A total of 1,730 publicly available genomes were sub-sampled, including 28 Guangdong genomes submitted by other institutions in 2025. Consequently, in addition to 1,263 ECSA sequences generated in this study, we included these 28 additional public genomes and assembled a final outbreak dataset of 1,291 genome isolates.

### Maximum likelihood analysis

To obtain a robust phylogenetic tree, we first removed potentially divergent genetic sequences. We used the RDP v5^49^ to detect evidence of potential recombination and excluded sequences where recombination was identified by at least six out of seven recombination detection methods^50–56^. We further removed genomes whose nucleotide composition significantly deviated from the average composition of the alignment using a composition chi-square test in IQ-TREE 2.4.0^57^.

We generated a preliminary maximum likelihood (ML) tree in IQ-TREE 2.4.0^57^ using the best-fit substitution model identified by ModelFinder^58^. We used TreeTime^59^ and TempEst^60^ to identify and remove temporal outliers. The curated datasets of outbreak clade-associated genomes alongside the background genomes showed sufficient temporal signal (**Fig. 2c**). The ML tree was re-estimated under the same conditions described above, and its branch support was inferred using an ultrafast bootstrap with 1,000 replicates.

### Discrete phylogeographic analysis

The initial ML phylogeny showed that CHIKV genomes from the Guangdong outbreak comprised two major clades (referred to as clade I and clade II, **Fig. 2a**). Hence, 99.5% (1,284/1,291) of Guangdong CHIKV genomes belonged to clade I, which became our focus of phylogeographic reconstruction.

To efficiently trace the geographical origin of the Guangdong outbreak, we only retained those phylogenetically close genomic sequences as background genomes. Specifically, all genomes from Central Africa and the Indian Ocean that fell within the ancestral clade of clade I were used. We discretized the locations of CHIKV sequences sampled outside Guangdong into three demes: Central Africa, Réunion, and other Indian Ocean countries (comprising Madagascar, Mauritius, and Mayotte). Similarly, we discretized sequence sampling locations in Guangdong into 12 geographic areas, including 9 cities (Foshan, Jiangmen, Zhuhai, Dongguan, Zhaoqing, Zhongshan, Guangzhou, Shenzhen, Huizhou) and 3 integrated sub-regions (West-Guangdong, North-Guangdong, and East-Guangdong) (**Fig. 1a**). The geographic integration of the last three sub-regions was due to the small number of sequences available, which can also accordingly reduce the number of demes and computational burden. Outbreak genomes were randomly sampled in proportion to the area-specific monthly case data to ensure that the genomic sample was representative of the underlying epidemic. To account for the possible overrepresentation of Réunion genomic sequences, we performed sensitivity analyses by reducing the number of genomes from Réunion (**Extended Data Fig. 4**).

Based on the curated dataset above, we performed the Bayesian phylogenetic and phylogeographic analyses using BEAST X v.10.5.0^61^ to infer time-calibrated phylogenies and ancestral geographic states along the phylogenies. The BEAGLE library v4 was used to accelerate its core calculations^62^. We employed a generalized time reversible (GTR) nucleotide substitution model with gamma-distributed among-site rate heterogeneity and invariant sites, as suggested by jModelTest v.2.1.10^63^. A previous methodological study also showed that Bayesian phylogenetic inference is robust to complex substitution models with well-behaved priors^64^. To evaluate the sensitivity of our findings to substitution-model specification, we repeated the analyses using an alternative nucleotide substitution model (GTR + I) and compared the tMRCA estimates and phylogeographic inferences (**Extended Data Fig. 6, Table S2**). We explored multiple combinations of molecular clock and coalescent models without informative priors and used path-sampling and stepping-stone approaches to select the best fitting model (**Extended Data Table 1**). The preferred model comprised an uncorrelated relaxed molecular clock with a lognormal distribution of branch rates^65^ and a Bayesian Skygrid tree prior. A prior with mean substitution rate of 5×10^-4^ substitutions per site per year (s/s/y) and a standard deviation (SD) of 1.5×10^-4^ s/s/y was specified. This corresponds to a 95% confidence interval of 2.06×10^-4^ to 7.94×10^-4^ s/s/y, which encompasses both previously reported CHIKV evolutionary rates^66,67^ and the posterior substitution rate obtained during the model selection runs (**Table S1**). We used a Bayesian Skygrid coalescent prior^68^, for which we specified grid points equidistantly spaced in 1-month interval for the most recent two years, and equidistantly spaced in 2-year interval for the past 50 years. For samples with incomplete dates, their dates were concurrently estimated assuming a uniform prior within the known temporal bounds. Phylogeographic reconstruction was conducted using the asymmetric discrete trait evolution model^69^ and the Bayesian stochastic search variable selection (BSSVS) approach. Bayesian analyses were run for 400 million Markov chain Monte Carlo (MCMC) steps, with states and trees sampled every 50,000 steps. Three independent MCMC chains were run and combined after discarding the first 10% as burn-in. Convergence of the analysis was assessed using Tracer 1.7.2^70^. Maximum clade credibility (MCC) trees were summarized using TreeAnnotator.

We estimated the intensity of Markov jump (i.e., number of transitions among each pair of locations) using TreeMarkovJumpHistoryAnalyzer tool^71^. Bayes factors (BF) were calculated to identify well-supported viral movement events, where > 100 was deemed as decisive support, 10 < BF ≤ 100 as strong support, and 3 < BF ≤ 10 as substantial support.

### Continuous phylogeographic analysis

To map the local spatiotemporal dynamic of outbreak clade I in Guangdong Province, we implemented a spatially explicit phylogeographic analysis using the continuous diffusion model in BEAST X v.10.5.0^61^. For 4.8% (n = 62) of sequences only geolocated to the city or district level, we randomly assigned latitude and longitude coordinates with sample weights proportional to the population density. To account for potential biases in genetic sampling efforts over space and time, we also performed a sub-sampling process to randomly select genomes in proportion to the city-specific monthly case data, which yielded a total of 661 genomes selected. For sequences originating from identical latitude and longitude coordinates, we added random jitter with noise drawn uniformly from a window size of 0.005 to resolve duplicated locations. To reduce computation burden and also account for uncertainty in the phylogenetic tree inference, analyses were conducted as described above using an uncorrelated relaxed lognormal molecular clock and the Bayesian Skygrid tree prior (informed by model selection, **Extended Data Table 2**) to obtain empirical trees. A total of 25 grids were set in Skygrid tree prior, each of which corresponds to weekly intervals. Because the outbreak genomes exhibited limited genetic diversity, we specified informative priors for both the tMRCA and substitution rate. For the tMRCA, we used a normally distributed prior with a mean of 0.48 years before the present (SD = 0.02), truncated to the interval 0.40–0.55 years (corresponding to 2 April to 27 May 2025). This interval was informed by the tMRCA estimates obtained from analyses incorporating background genomes (**Table S2**). For the mean substitution rate, we used a normally distributed prior with a mean of 5.88 × 10⁻⁴ substitutions per site per year (SD = 5 × 10⁻⁵), truncated to the interval 4.0 × 10⁻⁴ to 8.0 × 10⁻⁴ substitutions per site per year. This range was informed by substitution rate estimates obtained from the same analyses. Using Guangdong outbreak genomes alone, the strict and relaxed molecular-clock analyses produced nearly identical tMRCA estimates and posterior mean substitution rates: 3 May 2025 and 6.16 × 10⁻⁴ substitutions per site per year under the strict clock, compared with 4 May 2025 and 6.17 × 10⁻⁴ substitutions per site per year under the relaxed clock model (**Table S2**). The two clock models also produced consistent estimates of relative genetic diversity through time (**Fig. S4**).

Based on empirical trees, continuous phylogeographic reconstructions of the outbreak clade were conducted under a relaxed random walk diffusion model with branch-specific heterogeneity in diffusion velocity modelled using a Cauchy distribution^72^. Operator weights were assigned to tree distribution moves and location diffusion rates. The MCMC analysis was run for 100 million steps, with states and trees sampled every 50,000 steps.

The MCC tree was annotated by keeping node heights as in the selected tree after discarding the first 10% burn-in samples. The R package “seraphim” was used to visualize the continuous phylogeographic diffusion and estimate dispersal statistics^73^. Specifically, we estimated the maximal wavefront distances and the weighted diffusion coefficient (km^2^/year) by deriving branch-specific movement distances and times from spatially annotated phylogenetic nodes.

### Spatial net flow density analysis

We used the MCC tree from the continuous phylogeographic reconstruction to further perform a spatial net flow density analysis, similar to Tsui et al.^74^. Briefly, we summarized each dispersal event represented by a directed branch with a head node (origin) and a tail node (destination), along with their dates of occurrence. We only retained dispersal events with a geographic distance ≥ 5 km to remove short-range movements. By setting up a 200×200 grid over the study area, we performed Gaussian kernel density estimation (KDE) of the spatial distribution of origin (head nodes) and destinations (tail nodes) using a Gaussian kernel with bandwidth 0.4, respectively. We further computed the difference between the origin and destination density, and then re-scaled it to the range [-1,1], which was used to indicate whether there was a net flow of viral lineages into or out of the location.

### Sensitivity of clade I tMRCA estimates

Given that both the genome datasets and tree model might affect the estimates of internal node ages, we inferred the clade I tMRCA outbreak using multiple genomic datasets and approaches to test the robustness of our estimates. First, we extracted the tMRCA of the Guangdong genomes across posterior trees from the discrete phylogeographic analysis, in which we combined the Guangdong genomes with contextualized genomes from the Indian Ocean and Central Africa (dataset 1). Second, we assembled another genomic dataset containing only genomes from the Indian Ocean as background sequences (dataset 2). Third, instead of using any background genomes, we employed 661 local outbreak genomes alone that were selected for the continuous phylogeographic analysis (dataset 3) to infer the tMRCA. For each of these above three datasets, the phylogenetic inferences were performed using a relaxed clock model and the Skygrid tree prior. Fourth, we chose an episodic birth-death-sampling model^75^ to provide an additional estimate of the clade I tMRCA using dataset 3 under an alternative tree prior. Similar to Skygrid prior, we also set up 25 temporal epochs that correspond to weekly intervals in each. Birth and sampling rates are modelled as piecewise constant across epochs, while the death rate is assumed as a constant. We refer to the Gaussian Markov random fields (GMRF) smoothing prior settings on the birth, death, and sampling rates from previous work^76^. Specifically, the prior for the birth rate was calculated based on an empirical Bayes approach proposed by Magee et al.^77^; the constant death rate prior was obtained from an estimation of the infectious duration window covering 8 to 15 days; and the prior for sampling rate was set to reflect the current sampling fraction. Posterior distributions of the clade I tMRCA are shown in **Fig. 4a** and **Table S2**.

### Estimated importation intensity

To verify the robustness of the tMRCA estimates, we estimated the importation intensity from countries where ECSA lineages were circulating and for which data were available. The estimated importation intensity (EII) was calculated by multiplying the estimated monthly prevalence of ECSA lineage in each country and the number of air passengers arriving at Guangdong airports from that country. The monthly air passenger volumes were provided by Official Airline Guide (OAG) Ltd. through a data sharing agreement. The monthly prevalence of ECSA lineage was computed as the product of the CHIKV incidence and the proportion of CHIKV infections attributed to the ECSA lineage in that month. CHIKV case data at the monthly resolution were gathered from searches of the websites of the country’s ministry of health, local academic partner, official news, and published literature. The fraction of ECSA lineage was estimated from country-specific lineage percentages based on CHIKV genomes available in GISAID. When country-specific percentage data were not available, regional data were used instead. We note that EII dynamics should be regarded as a rough proxy rather than an accurate representation, because the metric accounts only for air-travel-based mobility and surveillance intensity may vary across space and time.

## Data Availability

All data produced in the present study are available upon reasonable request to the authors

## Acknowledgement

We gratefully acknowledge all data contributors, i.e., the authors and their originating laboratories responsible for obtaining the specimens, and their submitting laboratories for generating the genetic sequence and metadata and sharing via the GISAID Initiative, on which this research is based (https://doi.org/10.55876/gis8.260914ys). The computations in this research were performed using the CFFF platform of Fudan University. This work was supported by Guangdong Province S&T Program (2025B1111050002 to B.L., X.P., P.J., M.Z., and X.Z.), Guangdong Provincial Center for Disease Control and Prevention Supports Talent Program (2024D364 to B.L., C.Z., P.J., and Z.L.), the Talent Support Program of Guangdong Provincial Center for Disease Control and Prevention (2025D386 to H.Y.), the Prevention and Control of Emerging and Major Infectious Diseases-National Science and Technology Major Project (2025ZD01901300 to H.Y.), the National Natural Science Foundation of China (823B2089 to Z.C.), the Wellcome Trust Dengue and Zika Immunology and Genomics Multi-Country Network (DeZi Network) (316633/Z/24/Z to N.R.F.), the National Postdoctoral Program for Innovative Talent (BX20250185 to Z.C.), and the Young Scientists Fund of the National Natural Science Foundation of China (82304199 to J.C.).

## Author contributions

H.Y. conceived and supervised the research. Z.C., B.L., and H.Y. designed the research. X.P., P.J., C.Z., M.Z., Y.Z., S.H., X.Z., Z.L., J.S., C.S., Q.Y., Q.T., Z.Z., L.Z., J.H., and B.L. undertook fieldwork and experiments. Z.C., A.S., and W.L. collated the data. Z.C. analysed the data. Z.C., J.C., J.L., N.R.F., B.L., and H.Y. advised on methodologies. Z.C. wrote the initial manuscript draft. N.R.F. and H.Y. critically revised the manuscript. All authors edited, read, and approved the manuscript.

## Extended data

**Extended Data Fig. 1.**
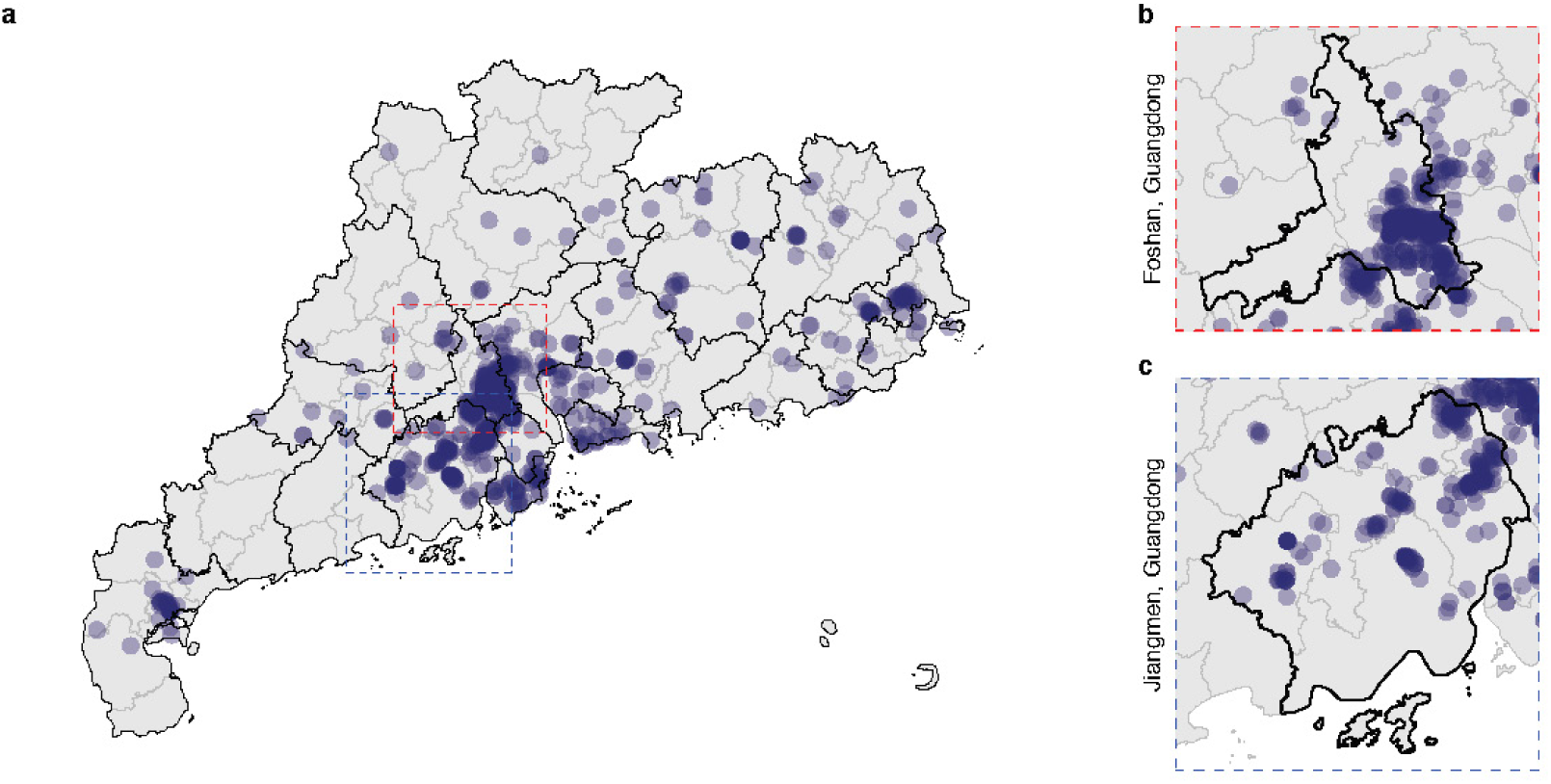
The geographic locations of CHIKV genomes generated in this study. The two rightmost panels (**b-c**) provide zoomed views of the genomic situation in Foshan and Jiangmen, Guangdong, respectively.

**Extended Data Fig. 2.**
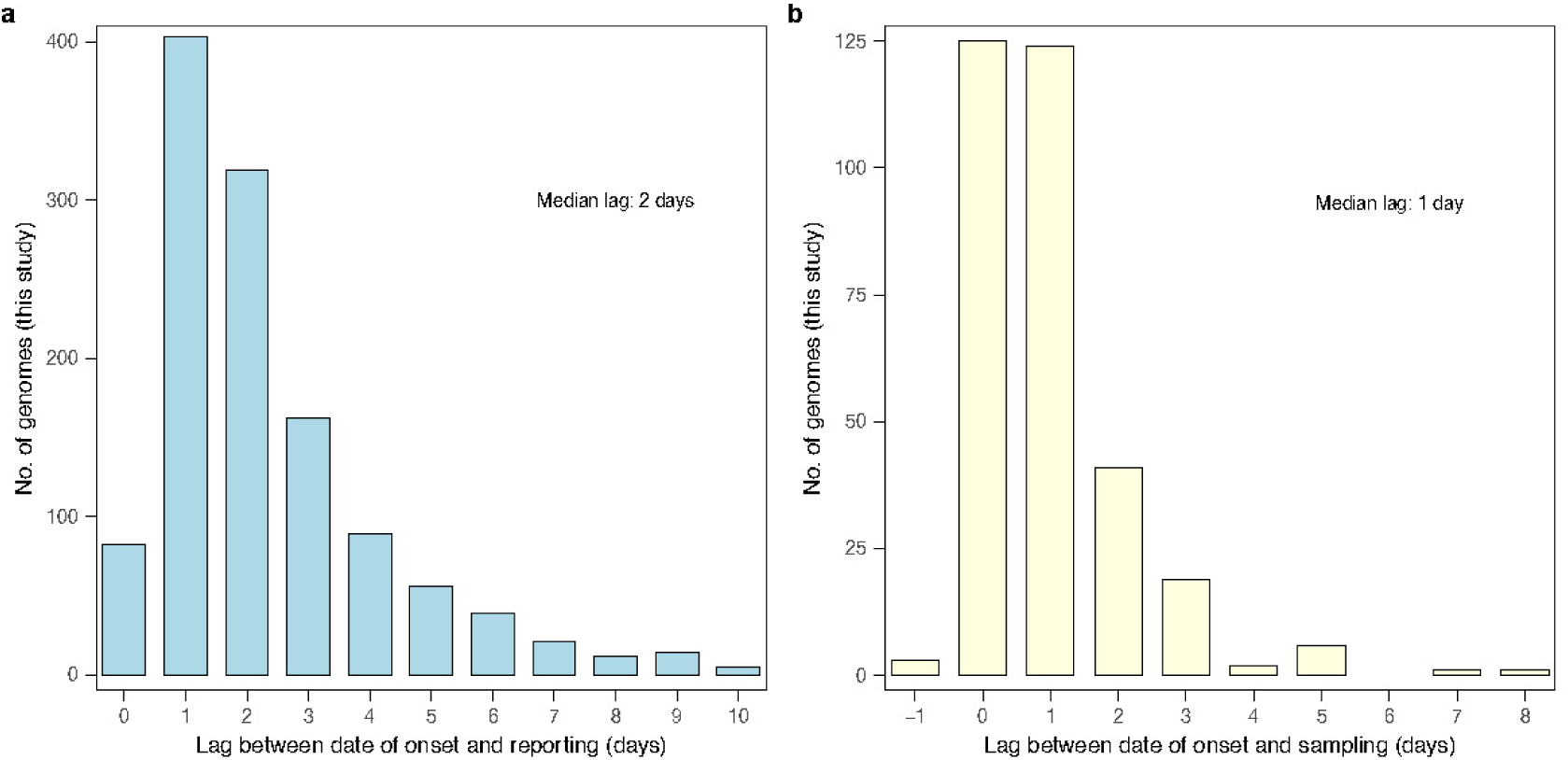
Intervals between the date of onset and the date of reporting/sample collection for the genomes generated in this study. (**a**) Intervals between the date of onset and the date of reporting. (**b**) Intervals between the date of onset and the date of sample collection. Two genomes were sampled before symptom onset.

**Extended Data Fig. 3.**
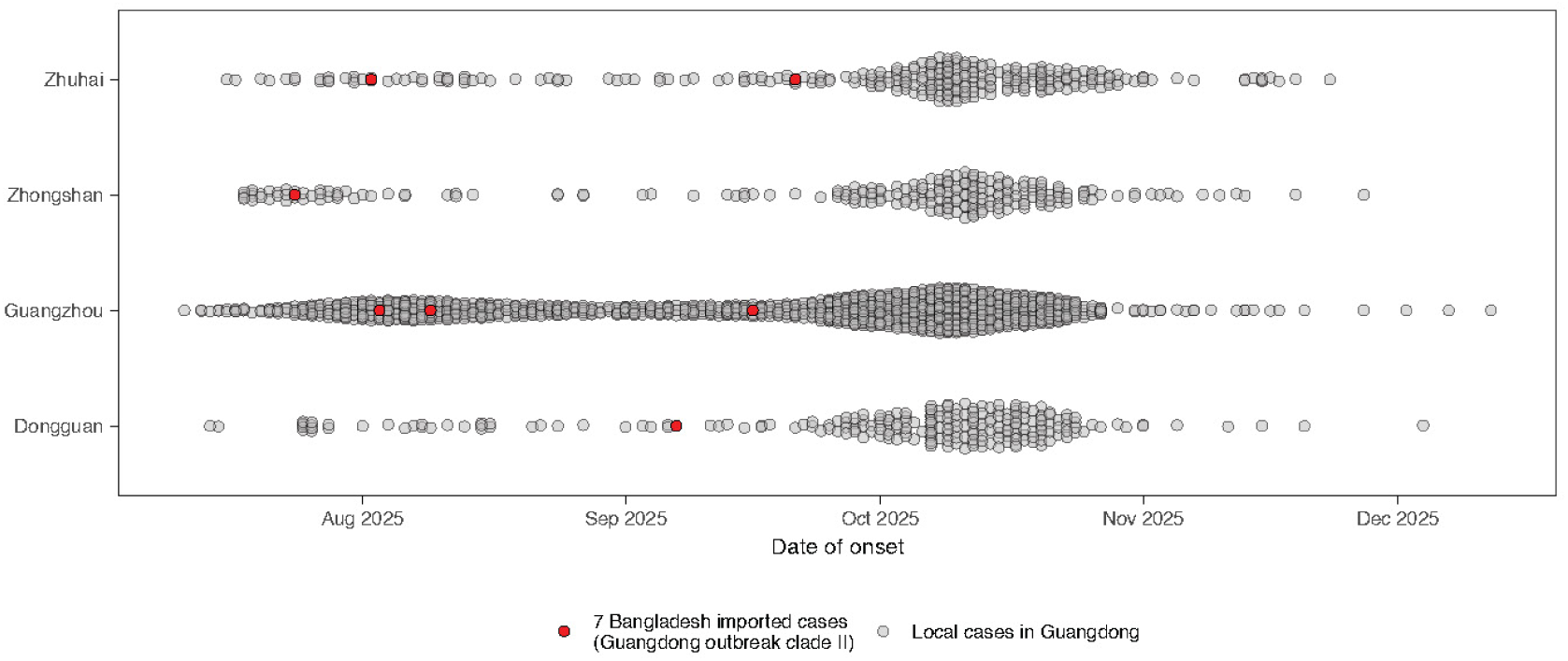
The temporal placement of seven CHIKV genomes belonging to outbreak clade II relative to the case occurrences. Gray points denote the local cases in Guangdong.

**Extended Data Fig. 4.**
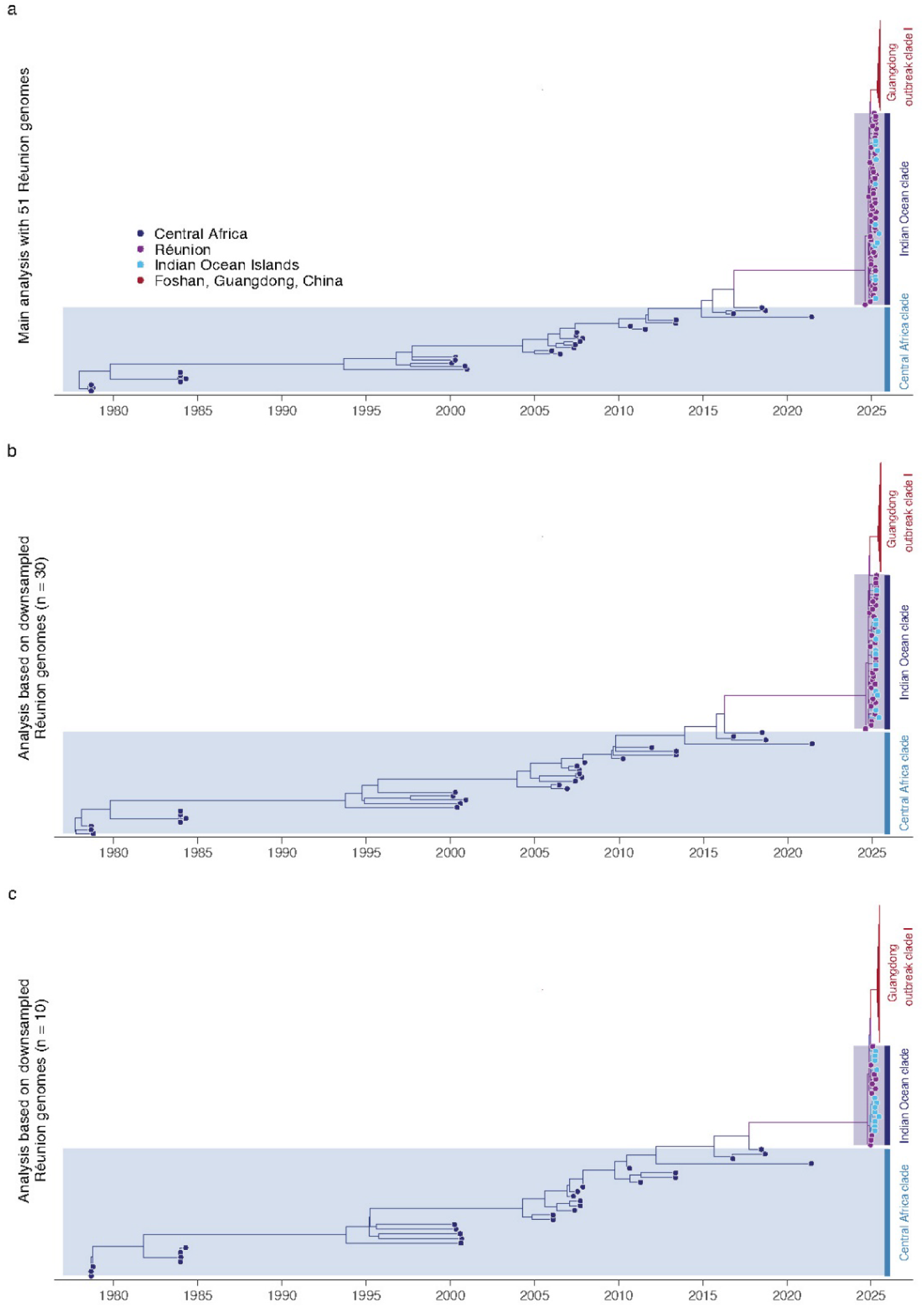
Maximum clade credibility phylogenies inferred under genomic datasets with reduced number of genomes from Réunion. (**a**) Main analysis with 51 genomes from Réunion. (**b**) Analysis based on downsampled Réunion genomes (n = 30). (**c**) Analysis based on downsampled Réunion genomes (n = 10). Tips are colored according to the sample locations and branches are colored by the inferred geographic states.

**Extended Data Fig. 5.**
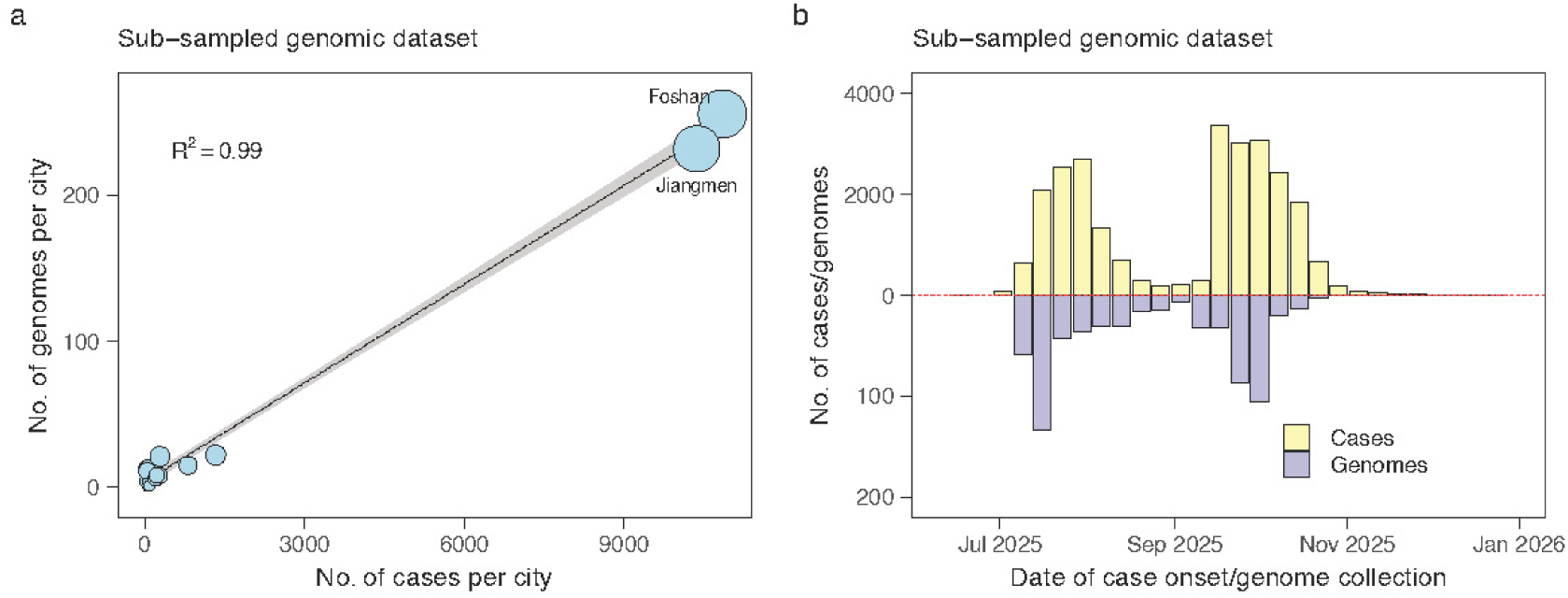
The spatiotemporal representative of CHIKV genomic dataset after the sub-sampling process. (**a**) The correlation between the number of CHIKV confirmed cases against the sub-sampled number of CHIKV genomes for each of the 21 prefecture-level cities in Guangdong. Circle sizes are proportional to the sub-sampled number of genomes. (**b**) The temporal distribution of genomes and cases after the sub-sampling process.

**Extended Data Fig. 6.**
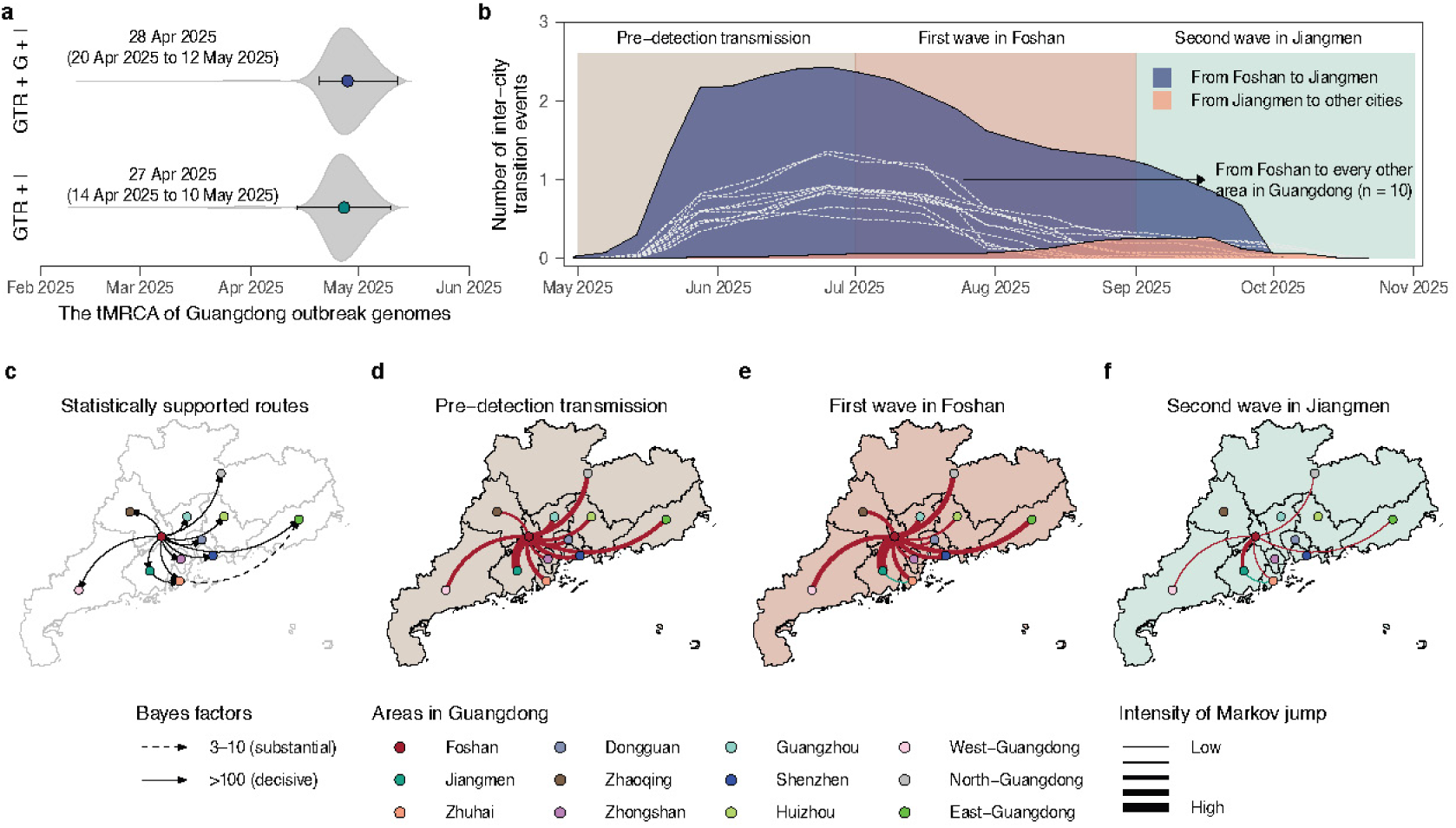
Sensitivity analyses assessing the impact of the GTR + I substitution model on clade I tMRCA estimates and phylogeographic inference. (**a**) Estimated posterior distributions of the tMRCA of the Guangdong outbreak genomes using the GTR + Γ + I and GTR + I substitution model, respectively. Points and ranges represent the posterior mean and 95% HPD intervals, respectively. (**b**) Weekly inter-city Markov jump counts (location transition events) through time based on GTR + I substitution model. (**c**) Migration routes with statistically supported evidence. (**d-f**) Estimated number of location transition events per period between each pair of geographic areas. Line thickness denotes the intensity of Markov jumps. GTR + Γ + I, a generalized time reversible (GTR) nucleotide substitution model with gamma-distributed among-site rate heterogeneity and invariant sites; GTR + I, a GTR substitution model with invariant sites.

**Extended Data Table 1.** Log marginal likelihood under different molecular clock and coalescent model combinations. The genomic datasets analyzed here were composed of Guangdong outbreak sequences together with phylogenetically close background sequences, which were used for discrete phylogeographic analysis. We tested three coalescent models: a constant population, an exponential growth population, and a nonparametric Bayesian Skygrid tree prior. We tested two molecular clock models: a strict molecular clock (Strict) and an uncorrelated relaxed molecular clock with lognormal distribution (UCLN). The best fitting model combination is ranked 1 (bold). PS, path sampling model selection, SS, stepping-stone model selection.

| **Model Combination** | **PS** | **PS-Ranking** | **SS** | **SS-Ranking** |
|---|---|---|---|---|
| Strict, Constant | -25061.447 | 6 | -25062.782 | 6 |
| Strict, Exponential | -25059.346 | 5 | -25060.855 | 5 |
| Strict, Skygrid | -24846.944 | 2 | -24849.091 | 2 |
| UCLN, Constant | -25048.057 | 3 | -25049.664 | 3 |
| UCLN, Exponential | -25049.589 | 4 | -25051.387 | 4 |
| **UCLN, Skygrid** | **-24834.611** | **1** | **-24836.947** | **1** |

**Extended Data Table 2.**
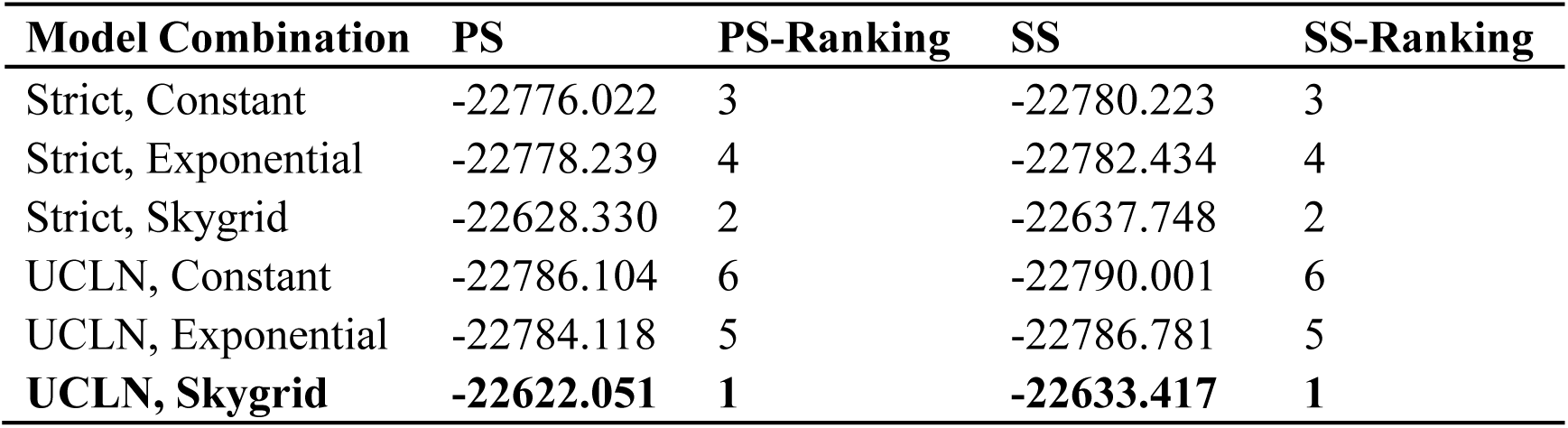
Log marginal likelihood under different molecular clock and coalescent model combinations. The genomic datasets analyzed here were Guangdong outbreak sequences alone, which were used for continuous phylogeographic analysis. We tested three coalescent models: a constant population, an exponential growth population, and a nonparametric Bayesian Skygrid tree prior. We tested two molecular clock models: a strict molecular clock (Strict) and an uncorrelated relaxed molecular clock with lognormal distribution (UCLN). The best fitting model combination is ranked 1 (bold). PS, path sampling model selection, SS, stepping-stone model selection.

## References

1 Weaver, S. C. & Lecuit, M. Chikungunya virus and the global spread of a mosquito-borne disease. The New England journal of medicine 372, 1231–1239, doi:10.1056/NEJMra1406035 (2015).

2 Chen, R. et al. Lineage Divergence and Vector-Specific Adaptation Have Driven Chikungunya Virus onto Multiple Adaptive Landscapes. mBio 12, e0273821, doi:10.1128/mBio.02738-21 (2021).

3 Ribeiro Dos Santos, G., et al. Global burden of chikungunya virus infections and the potential benefit of vaccination campaigns. Nature medicine 31, 2342–2349, doi:10.1038/s41591-025-03703-w (2025).

4 Farooq, Z. et al. Impact of climate and Aedes albopictus establishment on dengue and chikungunya outbreaks in Europe: a time-to-event analysis. The Lancet Planetary Health 9, e374–e383, doi:10.1016/S2542-5196(25)00059-2 (2025).

5 Menegale, F. et al. Risk assessment and perspectives of local transmission of chikungunya and dengue in Italy, a European forerunner. Nature communications 16, 6237, doi:10.1038/s41467-025-61109-1 (2025).

6 Wu, D. et al. Chikungunya outbreak in Guangdong Province, China, 2010. Emerging infectious diseases 18, 493–495, doi:10.3201/eid1803.110034 (2012).

7 Yang, Y. F. et al. The ecological factors contributing to the chikungunya outbreak in Ruili, a border city in Yunnan Province, China. BMC infectious diseases 25, 1643, doi:10.1186/s12879-025-12056-1 (2025).

8 Lu, W. et al. Estimating the epidemic size of chikungunya virus infection in Guangzhou, China, from July to September 2025: a single-center cross-sectional study. Emerging microbes & infections 15, 2645833, doi:10.1080/22221751.2026.2645833 (2026).

9 Sun, C. et al. Isolation and genomic characterization of Chikungunya virus during the 2025 outbreak in Foshan, China. Emerging microbes & infections 14, 2595790, doi:10.1080/22221751.2025.2595790 (2025).

10 Liang, J. et al. Emerging local chikungunya virus transmission in a major urban area in Southern China: characteristics of clinical manifestations, viral evolution and climatic influences. National science review 13, doi:10.1093/nsr/nwaf529 (2026).

11 Gong, W. et al. Outbreak of Chikungunya Virus with Aedes albopictus-Adaptive Mutations - Guangdong Province, China, 2025. China CDC Wkly 7, 1528–1532, doi:10.46234/ccdcw2025.260 (2025).

12 Cao, L. et al. Genomic characterization of a large-scale chikungunya outbreak in China. The Journal of infection, 106801, doi:10.1016/j.jinf.2026.106801 (2026).

13 Zhou, X. et al. First whole-genome chikungunya virus sequence detected in mosquitoes during the 2025 Foshan outbreak: evidence of field vector infection and transmission potential in China. Infectious diseases of poverty 14, 108, doi:10.1186/s40249-025-01383-9 (2025).

14 Yue, Y., Liu, Q., Liu, X., Zhao, N. & Yin, W. Dengue Fever in Mainland China, 2005-2020: A Descriptive Analysis of Dengue Cases and Aedes Data. International journal of environmental research and public health 19, doi:10.3390/ijerph19073910 (2022).

15 Chang, W. et al. Genomic epidemiology reveals the origins and transmission dynamics of chikungunya virus in China. Infectious diseases of poverty 15, doi:10.1186/s40249-026-01465-2 (2026).

16 Hill, V., Dellicour, S., Giovanetti, M. & Grubaugh, N. D. Phylogenetic insights into the transmission dynamics of arthropod-borne viruses. Nature Reviews Genetics, doi:10.1038/s41576-025-00854-x (2025).

17 Jia, P. et al. Comparative Analysis of Next- and Third-Generation Sequencing Platforms for Chikungunya Virus Whole-Genome Sequencing Using a Lineage-Inclusive Primer Set During the 2025 Foshan Outbreak. Tropical medicine and infectious disease 11, doi:10.3390/tropicalmed11020044 (2026).

18 Frumence, E. et al. Genomic insights into the re-emergence of chikungunya virus on Réunion Island, France, 2024 to 2025. Euro surveillance 30, doi:10.2807/1560-7917.Es.2025.30.22.2500344 (2025).

19 Tsetsarkin, K. A. & Weaver, S. C. Sequential adaptive mutations enhance efficient vector switching by Chikungunya virus and its epidemic emergence. PLoS pathogens 7, e1002412, doi:10.1371/journal.ppat.1002412 (2011).

20 Tsetsarkin, K. A. et al. Multi-peaked adaptive landscape for chikungunya virus evolution predicts continued fitness optimization in Aedes albopictus mosquitoes. Nature communications 5, 4084, doi:10.1038/ncomms5084 (2014).

21 Copernicus Climate Change Service, C. D. S. ERA5 post-processed daily-statistics on single levels from 1940 to present., 2024).

22 Huang, J. et al. The cross-city mobility patterns and incubation period of the early-stage chikungunya fever outbreak in Guangdong Province, China, in 2025. PLoS neglected tropical diseases 20, e0014360, doi:10.1371/journal.pntd.0014360 (2026).

23 Oliveira, T. d., et al. Reemergence of chikungunya in Mauritius driven by a novel lineage with pandemic potential. Research Square, doi:10.21203/rs.3.rs-8288410/v1 (2025).

24 Oidtman, R. J. et al. Inter-annual variation in seasonal dengue epidemics driven by multiple interacting factors in Guangzhou, China. Nature communications 10, 1148, doi:10.1038/s41467-019-09035-x (2019).

25 Meyer, A. D. et al. Predictability of infectious disease outbreak severity: Chikungunya as a case study. Science advances 11, eadt5419, doi:10.1126/sciadv.adt5419 (2025).

26 Grubaugh, N. D., Faria, N. R., Andersen, K. G. & Pybus, O. G. Genomic Insights into Zika Virus Emergence and Spread. Cell 172, 1160–1162, doi:10.1016/j.cell.2018.02.027 (2018).

27 Bustos Carrillo, F., et al. Epidemiological Evidence for Lineage-Specific Differences in the Risk of Inapparent Chikungunya Virus Infection. Journal of virology 93, doi:10.1128/jvi.01622-18 (2019).

28 Madewell, Z. J. et al. Leveraging Sentinel Enhanced Surveillance to Characterize the 2024-2025 Dengue Epidemic in Puerto Rico: Insights and Comparisons With Passive Surveillance. Public health reports (Washington, D.C. : 1974), 333549261440198, doi:10.1177/00333549261440198 (2026).

29 Hernandez-Romieu, A. C. et al. Utility of sentinel surveillance as an early warning system for emerging pathogens in a low-resource setting: Correlations of dengue, chikungunya, Zika, and COVID-19 trends between sentinel and passive surveillance systems in Puerto Rico, 2012-2023. PLOS Glob Public Health 6, e0006285, doi:10.1371/journal.pgph.0006285 (2026).

30 Faria, N. R. et al. Zika virus in the Americas: Early epidemiological and genetic findings. Science 352, 345–349, doi:10.1126/science.aaf5036 (2016).

31 Faria, N. R. et al. Genomic and epidemiological monitoring of yellow fever virus transmission potential. Science 361, 894–899, doi:10.1126/science.aat7115 (2018).

32 Liu, H., Zheng, W., Holmes, E. C. & Shi, W. Spatiotemporal dynamics of the unprecedented chikungunya outbreak in Guangdong, China. Emerging microbes & infections 15, 2695536, doi:10.1080/22221751.2026.2695536 (2026).

33 Cortes-Azuero, O. et al. The epidemiology of chikungunya virus in Brazil and the potential impact of vaccines: a mathematical modelling study. The Lancet. Infectious diseases, doi:10.1016/s1473-3099(25)00605-x (2025).

34 de Souza, W. M. et al. Chikungunya: a decade of burden in the Americas. *Lancet regional health*. Americas 30, 100673, doi:10.1016/j.lana.2023.100673 (2024).

35 Sharif, N., Khan, A., Sharif, N. & Dey, S. K. Spatiotemporal dynamics and reemergence of chikungunya virus in Bangladesh from 2008 to 2025. International journal of infectious diseases 166, 108529, doi:10.1016/j.ijid.2026.108529 (2026).

36 Zhang, M. et al. Epidemiological characteristics and transmission dynamics of the early stage Chikungunya fever outbreak in Foshan City, Guangdong Province, China in 2025. Infectious diseases of poverty 14, 93, doi:10.1186/s40249-025-01364-y (2025).

37 Clarke, J., et al. A global dataset of publicly available dengue case count data. Sci Data 11, 296, doi:10.1038/s41597-024-03120-7 (2024).

38 World Health Organization. WHO Global Dengue Dashboard, <https://worldhealthorg.shinyapps.io/dengue_global/> (2026).

39 Salje, H. et al. Reconstructing unseen transmission events to infer dengue dynamics from viral sequences. Nature communications 12, 1810, doi:10.1038/s41467-021-21888-9 (2021).

40 Lim, A. et al. The overlapping global distribution of dengue, chikungunya, Zika and yellow fever. Nature communications 16, 3418, doi:10.1038/s41467-025-58609-5 (2025).

41 World Health Organization. Policy considerations for strengthening preparedness and response to arbovirus epidemics and pandemics. (2025).

42 Chen, S. fastp 1.0: An ultra-fast all-round tool for FASTQ data quality control and preprocessing. iMeta 4, e70078, doi:10.1002/imt2.70078 (2025).

43 Danecek, P. et al. Twelve years of SAMtools and BCFtools. GigaScience 10, doi:10.1093/gigascience/giab008 (2021).

44 Grubaugh, N. D. et al. An amplicon-based sequencing framework for accurately measuring intrahost virus diversity using PrimalSeq and iVar. Genome biology 20, 8, doi:10.1186/s13059-018-1618-7 (2019).

45 Wallau, G. L. Arbovirus researchers unite: expanding genomic surveillance for an urgent global need. The Lancet. Global health 11, e1501–e1502, doi:10.1016/s2214-109x(23)00325-x (2023).

46 Zhang, Y. et al. Genomic characterization and evolutionary analysis of Chikungunya virus strains in Guangzhou, China, 2025. Virology journal 23, 15, doi:10.1186/s12985-025-03051-8 (2025).

47 Vilsker, M. et al. Genome Detective: an automated system for virus identification from high-throughput sequencing data. Bioinformatics 35, 871–873, doi:10.1093/bioinformatics/bty695 (2019).

48 Katoh, K. & Standley, D. M. MAFFT Multiple Sequence Alignment Software Version 7: Improvements in Performance and Usability. Molecular Biology and Evolution 30, 772–780, doi:10.1093/molbev/mst010 %J Molecular Biology and Evolution (2013).

49 Martin, D. P., et al. RDP5: a computer program for analyzing recombination in, and removing signals of recombination from, nucleotide sequence datasets. Virus evolution 7, veaa087, doi:10.1093/ve/veaa087 (2021).

50 Martin, D. & Rybicki, E. RDP: detection of recombination amongst aligned sequences. Bioinformatics 16, 562–563, doi:10.1093/bioinformatics/16.6.562 (2000).

51 Padidam, M., Sawyer, S. & Fauquet, C. M. Possible emergence of new geminiviruses by frequent recombination. Virology 265, 218–225, doi:10.1006/viro.1999.0056 (1999).

52 Martin, D. P., Posada, D., Crandall, K. A. & Williamson, C. A modified bootscan algorithm for automated identification of recombinant sequences and recombination breakpoints. AIDS research and human retroviruses 21, 98–102, doi:10.1089/aid.2005.21.98 (2005).

53 Smith, J. M. Analyzing the mosaic structure of genes. Journal of molecular evolution 34, 126–129, doi:10.1007/bf00182389 (1992).

54 Posada, D. & Crandall, K. A. Evaluation of methods for detecting recombination from DNA sequences: computer simulations. Proceedings of the National Academy of Sciences of the United States of America 98, 13757–13762, doi:10.1073/pnas.241370698 (2001).

55 Gibbs, M. J., Armstrong, J. S. & Gibbs, A. J. Sister-scanning: a Monte Carlo procedure for assessing signals in recombinant sequences. Bioinformatics 16, 573–582, doi:10.1093/bioinformatics/16.7.573 (2000).

56 Lam, H. M., Ratmann, O. & Boni, M. F. Improved Algorithmic Complexity for the 3SEQ Recombination Detection Algorithm. Mol Biol Evol 35, 247–251, doi:10.1093/molbev/msx263 (2018).

57 Minh, B. Q. et al. IQ-TREE 2: New Models and Efficient Methods for Phylogenetic Inference in the Genomic Era. Mol Biol Evol 37, 1530–1534, doi:10.1093/molbev/msaa015 (2020).

58 Kalyaanamoorthy, S., Minh, B. Q., Wong, T. K. F., von Haeseler, A. & Jermiin, L. S. ModelFinder: fast model selection for accurate phylogenetic estimates. Nat Methods 14, 587–589, doi:10.1038/nmeth.4285 (2017).

59 Sagulenko, P., Puller, V. & Neher, R. A. TreeTime: Maximum-likelihood phylodynamic analysis. Virus evolution 4, vex042, doi:10.1093/ve/vex042 (2018).

60 Rambaut, A., Lam, T. T., Max Carvalho, L. & Pybus, O. G. Exploring the temporal structure of heterochronous sequences using TempEst (formerly Path-O-Gen). Virus evolution 2, vew007, doi:10.1093/ve/vew007 (2016).

61 Baele, G. et al. BEAST X for Bayesian phylogenetic, phylogeographic and phylodynamic inference. Nat Methods, doi:10.1038/s41592-025-02751-x (2025).

62 Ayres, D. L. et al. BEAGLE: an application programming interface and high-performance computing library for statistical phylogenetics. Systematic biology 61, 170–173, doi:10.1093/sysbio/syr100 (2012).

63 Darriba, D., Taboada, G. L., Doallo, R. & Posada, D. jModelTest 2: more models, new heuristics and parallel computing. Nat Methods 9, 772, doi:10.1038/nmeth.2109 (2012).

64 Guimarães Fabreti, L. & Höhna, S. Nucleotide Substitution Model Selection Is Not Necessary for Bayesian Inference of Phylogeny With Well-Behaved Priors. Systematic biology 72, 1418–1432, doi:10.1093/sysbio/syad041 (2023).

65 Drummond, A. J., Ho, S. Y., Phillips, M. J. & Rambaut, A. Relaxed phylogenetics and dating with confidence. PLoS biology 4, e88, doi:10.1371/journal.pbio.0040088 (2006).

66 Volk, S. M. et al. Genome-scale phylogenetic analyses of chikungunya virus reveal independent emergences of recent epidemics and various evolutionary rates. Journal of virology 84, 6497–6504, doi:10.1128/jvi.01603-09 (2010).

67 Krambrich, J. et al. The evolutionary and molecular history of a chikungunya virus outbreak lineage. PLoS neglected tropical diseases 18, e0012349, doi:10.1371/journal.pntd.0012349 (2024).

68 Gill, M. S. et al. Improving Bayesian population dynamics inference: a coalescent-based model for multiple loci. Mol Biol Evol 30, 713–724, doi:10.1093/molbev/mss265 (2013).

69 Lemey, P., Rambaut, A., Drummond, A. J. & Suchard, M. A. Bayesian phylogeography finds its roots. PLoS computational biology 5, e1000520, doi:10.1371/journal.pcbi.1000520 (2009).

70 Rambaut, A., Drummond, A. J., Xie, D., Baele, G. & Suchard, M. A. Posterior Summarization in Bayesian Phylogenetics Using Tracer 1.7. Systematic biology 67, 901–904, doi:10.1093/sysbio/syy032 %J Systematic Biology (2018).

71 Lemey, P. et al. Untangling introductions and persistence in COVID-19 resurgence in Europe. Nature 595, 713–717, doi:10.1038/s41586-021-03754-2 (2021).

72 Lemey, P., Rambaut, A., Welch, J. J. & Suchard, M. A. Phylogeography takes a relaxed random walk in continuous space and time. Mol Biol Evol 27, 1877–1885, doi:10.1093/molbev/msq067 (2010).

73 Dellicour, S., Faria, N. R., Rose, R., Lemey, P. & Pybus, O. G. SERAPHIM 2.0: an extended toolbox for studying phylogenetically informed movements. Bioinformatics 42, btag093, doi:10.1093/bioinformatics/btag093 (2026).

74 Tsui, J. L. et al. Genomic assessment of invasion dynamics of SARS-CoV-2 Omicron BA.1. Science 381, 336–343, doi:10.1126/science.adg6605 (2023).

75 Shao, Y., Magee, A. F., Vasylyeva, T. I. & Suchard, M. A. Scalable gradients enable Hamiltonian Monte Carlo sampling for phylodynamic inference under episodic birth-death-sampling models. PLoS computational biology 20, e1011640, doi:10.1371/journal.pcbi.1011640 (2024).

76 Frumence, E. et al. Unravelling the epidemiological and dispersal dynamics of the 2024-2025 chikungunya virus outbreak on Réunion island. medRxiv, 2026.2001.2007.26343606, doi:10.64898/2026.01.07.26343606 (2026).

77 Magee, A. F., Höhna, S., Vasylyeva, T. I., Leaché, A. D. & Minin, V. N. Locally adaptive Bayesian birth-death model successfully detects slow and rapid rate shifts. PLoS computational biology 16, e1007999, doi:10.1371/journal.pcbi.1007999 (2020).

