## Supplementary material for "Genomic epidemiology reconstructs the pre-detection transmission and establishment of chikungunya virus in Guangdong, China": SI

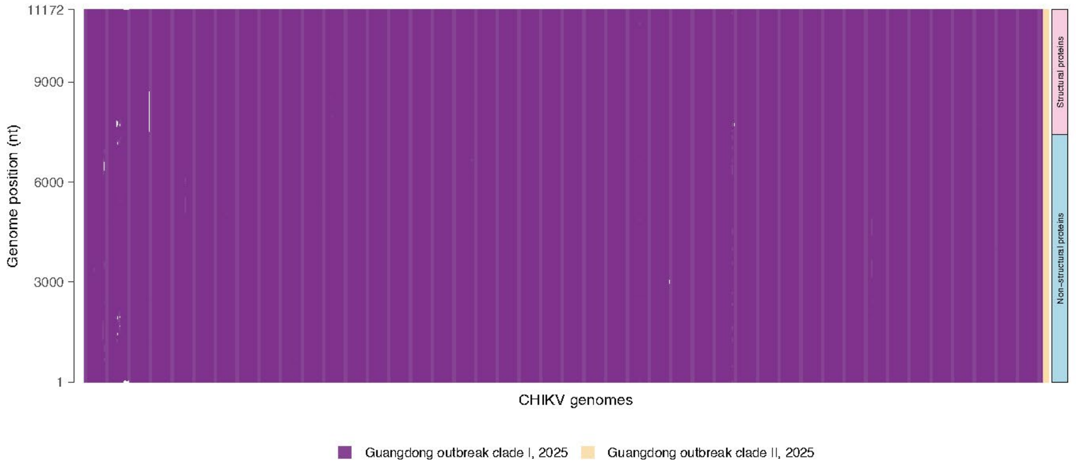


**Figure S1. The sequencing coverage across the CHIKV genomes generated in this study.** The bars are colored by the clade type, with white representing missing or ambiguous genome positions.

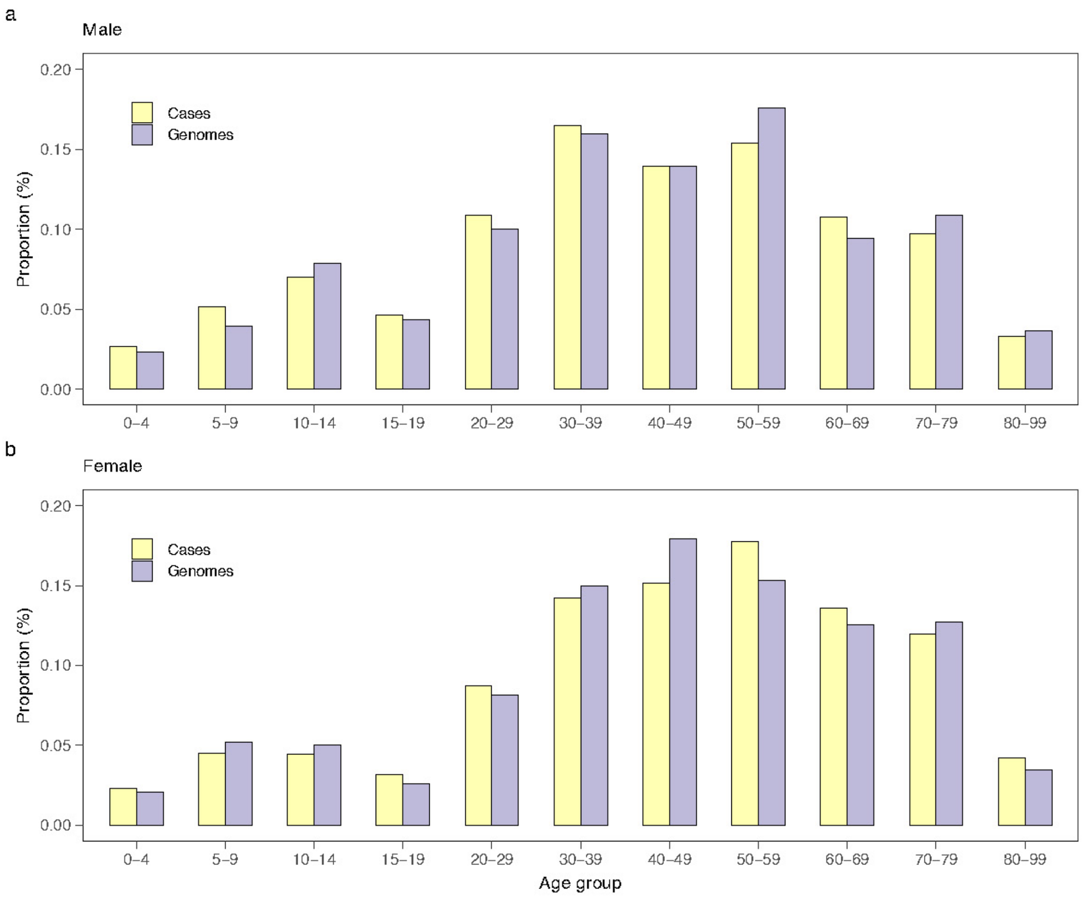


**Figure S2. The age and sex distribution of CHIKV cases and genomes in the 2025 Guangdong outbreak.** (**a**) The age-specific proportion of cases and genomes in males. (**b**) The age-specific proportion of cases and genomes in females.

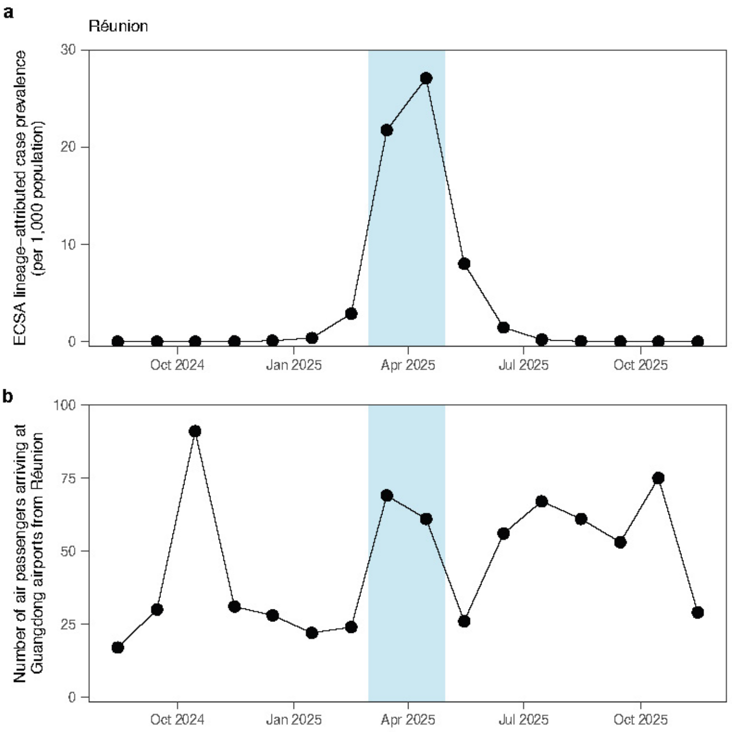


**Figure S3. Elements in calculating the estimated importation intensity (EII) in Réunion.** (**a**) Local ECSA lineage-attributed case prevalence in Réunion. (**b**) Number of air passengers arriving at Guangdong airports from Réunion over time.

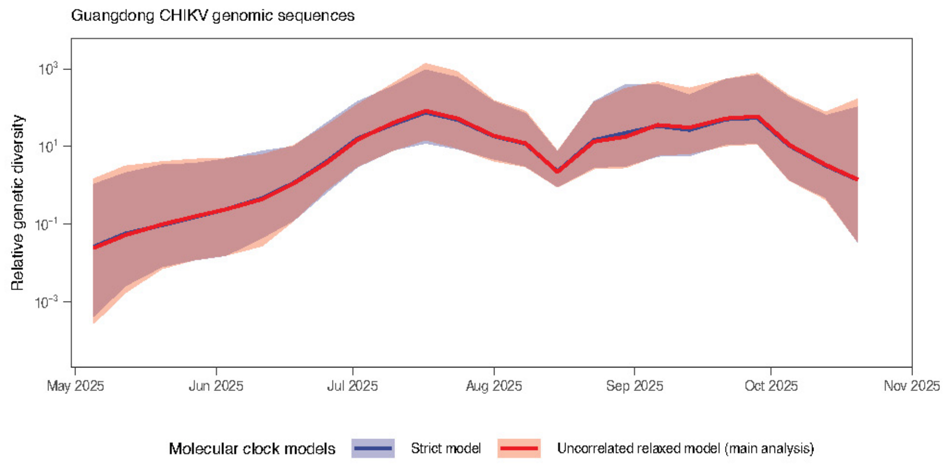


**Figure S4. Relative genetic diversity of chikungunya viruses in Guangdong.** The results were inferred by Bayesian Skygrid population analysis of a total of 661 Guangdong outbreak genomes, in which a strict molecular clock (blue line; visually difficult to distinguish due to the overlap of the two curves) and an uncorrelated relaxed molecular clock model (red line) was used, respectively.

**Table S1. Posterior estimates of the CHIKV substitution rate (substitutions per site per year) under different molecular clock models and coalescent models**. The genomic datasets analyzed here were composed of Guangdong outbreak sequences together with phylogenetically close background sequences, which were used for discrete phylogeographic analysis. Here we set the molecular clock rate to follow a non-informative continuous time Markov chain (CTMC) rate prior. HPD, highest posterior density. The mean substitution rate (mean of the rates of each branch weighted by the time length of the branch) was reported for a relaxed clock model.

| **Clock model** | **Coalescent model** | **Estimated substitution rate** | | |
|---|---|---|---|---|
| | | **Mean** | **Lower 95% HPD** | **Upper 95% HPD** |
| Strict | Constant | 5.23E-4 | 4.37E-4 | 6.04E-4 |
| Strict | Exponential | 5.28E-4 | 4.39E-4 | 6.09E-4 |
| Strict | Skygrid | 4.63E-4 | 3.94E-4 | 5.34E-4 |
| UCLN | Constant | 6.90E-4 | 5.61E-4 | 8.24E-4 |
| UCLN | Exponential | 7.11E-4 | 5.82E-4 | 8.47E-4 |
| UCLN | Skygrid | 4.95E-4 | 4.12E-4 | 5.79E-4 |

**Table S2. Sensitivity of the estimated time of the most recent common ancestor of Guangdong outbreak clade I and its parent node to genomic dataset and model configuration.** GTR + Γ + I: generalized time reversible (GTR) nucleotide substitution model with gamma (Γ) distributed among-site rate heterogeneity and a proportion of invariant sites (I); GTR + I: GTR substitution model with a proportion of invariant sites; HPD: highest posterior density; tMRCA, time of the most recent common ancestor. Parent node dates are reported only for analyses containing background genomes, because the parent node of the Guangdong clade cannot be estimated from Guangdong outbreak genomes alone.

| Genomic datasets | Model configuration | Clade I tMRCA (mean, 95% HPD) | Parent node of the clade I MRCA, date (mean, 95% HPD) |
|---|---|---|---|
| Guangdong + Indian Ocean + Central Africa genomes (Dataset 1) | GTR + Γ + I; relaxed molecular clock; Skygrid prior | 28 Apr 2025 (20 Apr 2025 to 12 May 2025) | 27 Dec 2024 (31 Oct 2024 to 11 Apr 2025) |
| Guangdong + Indian Ocean + Central Africa genomes (Dataset 1) | GTR + I; relaxed molecular clock; Skygrid prior | 27 Apr 2025 (14 Apr 2025 to 10 May 2025) | 25 Dec 2024 (19 Oct 2024 to 3 Apr 2025) |
| Guangdong + Indian Ocean genomes (Dataset 2) | GTR + Γ + I; relaxed molecular clock; Skygrid prior | 4 May 2025 (18 Apr 2025 to 14 May 2025) | 25 Dec 2024 (25 Oct 2024 to 5 Apr 2025) |
| Guangdong genomes (Dataset 3) | GTR + Γ + I; relaxed molecular clock; Skygrid prior | 4 May 2025 (20 Apr 2025 to 18 May 2025) | - |
| Guangdong genomes (Dataset 3) | GTR + Γ + I; strict molecular clock; Skygrid prior | 3 May 2025 (19 Apr 2025 to 16 May 2025) | - |
| Guangdong genomes (Dataset 3) | GTR + Γ + I; relaxed molecular clock; episodic birth-death sampling model | 5 May 2025 (21 Apr 2025 to 18 May 2025) | - |
